# Factors associated with severe COVID-19 outcomes among adults with at least a primary vaccination schedule: a retrospective cohort study from Alberta, Canada

**DOI:** 10.1101/2025.08.19.25334000

**Authors:** Sylvia Aponte-Hao, Huong Luu, Karen J.B. Martins, Jason R. Randall, Lynora Saxinger, Elissa Rennert-May, Jenine Leal, Tyler Williamson, Amanda Wilson, Michelle Blake, Tianyi Lu, Shehzad M. Iqbal, James A. Mansi, Andre B. Araujo, David Martin, Scott W. Klarenbach

**Author notes:** Corresponding Author: Scott W. Klarenbach.

## Abstract

**Background:** With a large proportion of adults in Canada vaccinated against SARS-CoV-2, it is important to identify who remain at risk for severe COVID-19 outcomes after primary vaccination/additional (booster) doses.

**Methods:** Adults (≥18 years) who received at least a primary vaccination schedule between December 2020 and March 2023 in Alberta, Canada were identified through population-level administrative data. Factors associated with severe COVID-19 outcomes (COVID-19 related hospitalization or death) were examined using univariable and multivariable Cox proportional hazard models.

**Results:** Among those who received a primary vaccination schedule (n=2,639,731), 0.2% (n=5,475) had ≥1 severe COVID-19 outcome. Factors associated with a greater risk of a severe COVID-19 outcome included older age (≥65 versus <65 years; unadjusted hazard ratio [95% confidence interval]: 8.83 [8.33-9.35]), and living with a higher comorbid burden (severe versus no burden; 32.56 [29.78-35.60]) or health condition of interest (1.33 [1.25-1.42] to 29.25 [23.14-36.99]) such as the relatively common condition of cardiovascular disease (7.01 [6.62-7.42]; present in 10.5% of the cohort) and less common condition of dementia (13.96 [12.76-15.28]; present in 1.5% of the cohort); male sex was also associated with risk (versus female; 1.07 [1.01-1.13]). Results remained significant in fully adjusted analysis, and among those with ≥1 additional vaccination dose (n=1,580,110).

**Conclusion:** Although primary vaccination and additional doses have dramatically reduced the occurrence of severe COVID-19 outcomes, results showed that continued risk remained for specific groups. Findings can be used to inform decision making, public health strategies, and policy development to protect those who remain at risk of severe COVID-19 outcomes after receipt of a primary vaccination schedule or additional doses, and support Canada’s National Advisory Committee on Immunisation recommendations for specific groups to stay up to date with COVID-19 vaccination to protect against severe COVID-19 outcomes.

## 1. Introduction

The coronavirus disease 2019 (COVID-19) pandemic, caused by severe acute respiratory syndrome coronavirus 2 (SARS-CoV-2), has been the most widespread and disruptive public health crisis of the 21st century, with over 778 million identified cases and 7.1 million deaths reported worldwide (as of August 3, 2025) [1]. Vaccination against COVID-19 has been shown to be highly effective in reducing severe COVID-19 outcomes such as hospitalization, intensive care unit admission, and death [2, 3]; it has been crucial in controlling the pandemic, and remains a cornerstone public health intervention for mitigating severe COVID-19 outcomes during the endemic period [4]. However, COVID-19 vaccine effectiveness and immunity acquired through SARS-CoV-2 infection may wane over time. This attenuation can be influenced by host factors such as older age and underlying health conditions that impair the immune response, viral factors including the emergence of new variants, and the degree of match between vaccine or infection-induced immunity and circulating variants [5–8]. Additional (booster) doses of the COVID-19 vaccine have been shown to increase protection by further reducing the risk of infection, particularly against severe outcomes [8–10].

While the majority of people in Canada have completed a primary COVID-19 vaccination schedule (82.0%), and 49.6% have received at least one additional dose (as of August, 2022) [11], some individuals remain at high risk of severe COVID-19 outcomes, particularly older adults, and individuals living with a higher comorbid burden and certain health conditions [12–20]. Understanding factors associated with severe COVID-19 outcomes among people who have received at least a primary vaccination schedule can support optimisation of vaccination recommendations, treatment strategies, and health system planning. However, limited evidence exists in Canada. Addressing this knowledge gap can support vaccine immunisation advisory recommendations, inform individual and clinical decision making, guide effective implementation of public health measures, and contribute to ongoing evidence-based policy decisions in the long-term management of COVID-19. The objective of this study was to identify factors associated with severe COVID-19 outcomes among adults who received a primary vaccination schedule, and describe risk profiles among this population in Alberta, Canada; those who received ≥1 additional vaccination dose were also examined.

## 2. Methods

Ethics approval was received from the University of Alberta Research Ethics Board (Pro00135542) that granted an exemption from requiring written informed consent (a waiver of consent was applied). Data custodian approvals were received from Alberta Health and Alberta Health Services (AHS) for the use of administrative health data. This study was reported according to the Reporting of studies Conducted using Observational Routinely-collected health Data guidelines [21].

### 2.1. Study design

This real world retrospective, observational, population-level cohort study was conducted using administrative health data from Alberta, Canada between April 1, 2015 and March 31, 2023. It encompassed an inclusion period from December 14, 2020 (start of COVID-19 vaccination in Alberta) to March 17, 2023, a 5 year look-back period for the assessment of baseline characteristics, and a follow-up period up to March 31, 2023.

### 2.2. Data source

The Canadian health care system is a single-payer system that provides publicly funded medically necessary care for all residents; universal prescription drug coverage is not included. In Alberta, the fourth most populous province in Canada (4.4-4.7 million people with 3.4-3.7 adults between 2020 and 2023), healthcare is administered under the Alberta Health Care Insurance Plan (AHCIP), of which over 99% of Albertans participate [22, 23].

A person-level linked (using a unique individual identifier [Personal Health Number]) data extract was created from the following listed databases by the data custodians and provided to the researchers (analysis was completed within the secure data custodian environment). The Population Registry contains demographic information for all Alberta residents with AHCIP coverage; elements include migration in and out of the province, and birth and death indicators. The Immunization and Adverse Reactions to Immunizations database contains records for vaccinations within the province, including data from AHS and pharmacist immunizers. The Discharge Abstract Database and National Ambulatory Care Reporting System include information on all individuals discharged from hospitals and facility-based ambulatory care (includes publicly-funded emergency department visits, and non-emergent outpatient treatments and procedures including same-day surgery, day procedures, and community rehabilitation program services), respectively – most responsible and secondary diagnostic fields are available and use International Classification of Disease – Version 10 – Canadian Enhancement (ICD-10-CA) codes. The Practitioner Claims database includes information on physician billing – up to three ICD – Version 9 – Clinical Modification (ICD-9-CM; Alberta specific) diagnostic codes can be listed per visit. The Alberta Cancer Registry collects information on demographics, tumor characteristics, and primary treatment from all individuals who received treatment in the province at the time of their initial cancer diagnosis. The Alberta Continuing Care Information System contains information on residents living in long-term care facilities and designated supportive living. The Pharmaceutical Information Network contains information on all community-dispensed prescription medications from pharmacies (regardless of payer). The Laboratory Information System includes laboratory test results collected by AHS. The Vital Statistics database contains information on births and deaths reported to Alberta Health. Records that were duplicates or contained an invalid Personal Health Number were discarded by the data custodians. Variables were checked for missing data and inconsistencies by the researchers; inconsistent data were corrected using data logic or information majority.

### 2.3. Cohort selection

Eligibility criteria for the cohort included those who: 1) completed a primary vaccination schedule of 2 doses of BNT162b2 (Pfizer-BioNTech), mRNA-1273 (Moderna), and/or ChAdOx1 nCoV-19 (Oxford-AstraZeneca), 2 doses of Novavax Nuvaxoid COVID-19 vaccine (Novavax), or 1 dose of Ad26.COV2.S (Janssen) between December 14, 2020 and March 17, 2023, 2) were aged ≥18 years on the date of the first COVID-19 vaccine dose, and 3) had AHCIP coverage for ≥5 years before the date of completion of the primary vaccination schedule (index date).

Follow-up commenced 14 days after the index date to allow sufficient time for the development of a full immune response [24], until a severe COVID-19 outcome (COVID-19 related hospitalization or death), 7 days following the first additional COVID-19 vaccination dose, loss to follow-up (relocation out of the province, non-COVID-19 death), or the end of the observation period (March 31, 2023). Individuals were excluded if they had a severe COVID-19 outcome, COVID-19 related emergency department visit, a positive SARS-CoV-2 reverse transcription-polymerase chain reaction (RT-PCR) test result, or were lost to follow-up <14 days after the index date. A sub-group consisted of individuals within the cohort who had ≥1 additional COVID-19 vaccination dose; follow-up commenced 7 days after the first additional dose.

### 2.4. Study measures

Demographic characteristics recorded on the index date included age, sex, urban/rural residence (determined by the second digit of the postal code), and those residing within a long-term care facility. Socioeconomic status was measured using the Pampalon material deprivation index (MDI; based on education, employment status, and average income) [25]; components are derived from Canadian census data at the dissemination area level that is linkable to postal code, and presented based on quintiles from most well-off (quintile 1) to most deprived (quintile 5) that represent the Alberta general population.

Clinical characteristics included a measure of overall comorbid burden – the Charlson Comorbidity Index score – that was measured over a 2 year rolling window during the 5 year period before the index date (Supplementary Table 1) [26, 27], and health conditions of interest – asthma, cardiovascular disease, chronic kidney disease, chronic obstructive pulmonary disease, dementia, diabetes, Down’s syndrome, dyslipidemia, hypertension, an immunocompromised status (autoimmune disorders; cancer and receiving chemotherapy or an immunocompromising drug; history of solid organ or bone marrow transplant; receiving dialysis; and ‘other immunocompromising conditions’ that included diseases of the blood, human immunodeficiency virus, immune system disorders, sickle cell anaemia, spleen malformations or removal, and treated for tuberculosis), a mental health disorder (bipolar disorder, depression, or schizophrenia), obesity, Parkinson’s disease, pregnancy, and pulmonary hypertension (Supplementary Table 2). The proportion of those who had a prior SARS-CoV-2 infection (defined as ≥1 laboratory confirmed infection or COVID-19 related hospitalization or emergency department visit before the index date), and those who previously received a community pharmacy dispensation of nirmatrelvir-ritonavir were reported.

The primary outcome of interest was a severe COVID-19 outcome (COVID-19 related hospitalization or death) that occurred ≥14 days after the index date, or ≥7 days after the first additional dose [24, 28]. A COVID-19 related hospitalization was defined as a hospital admission with an ICD-10-CA U07.1 code located within any diagnostic field, U07.3 located in a secondary diagnostic field (algorithm previously validated in Alberta that resulted in high sensitivity [94.4%] and positive predictive value [94.2%] [29]), or J04, J09-J22 (except J19), J80, or U04 located in the most responsible diagnostic field and a positive SARS-CoV-2 RT-PCR test result ≤14 days prior to or during the hospital admission (Supplementary Table 3 lists code descriptions) [30, 31]; those who had post-admission acquired COVID-19 were excluded. COVID-19 related mortality was defined as death during a COVID-19 related hospitalization, an all-cause death ≤28 days after a positive SARS-CoV-2 RT-PCR test result, or COVID-19 listed as the primary reason for death in the vital statistics record.

### 2.5. Statistical analysis

Descriptive statistics included counts and percentages for categorical variables, and means and standard deviations (SD) or medians and interquartile ranges (Q1-Q3) for continuous variables, as appropriate. Cumulative 1-year incidence rates and incidence density rates, and probabilities at 6-, 12-, and 24-months (with 95% confidence intervals [95% CI]) after the index date were calculated among the primary vaccination schedule cohort, and presented by factor according to the type of severe COVID-19 outcome event (any severe COVID-19 outcome; hospitalization [non-intensive care unit], intensive care unit admission, death).

Univariable and multivariable Cox proportional hazards models were used to determine relative differences in the hazards (unadjusted and adjusted hazard ratios [uHR, aHR] with 95% confidence intervals [CI]) of severe COVID-19 outcomes (any severe COVID-19 outcome; hospitalization [non-intensive care unit], intensive care unit admission, death) between individuals with and without specific factors. Covariates for multivariable models were selected based on a pre-specified conceptual framework using a directed acyclic graph [32]. The full adjustment set included age, sex, urban/rural and long-term care facility residence, socioeconomic status (MDI), overall comorbid burden (Charlson Comorbidity Index score), cardiovascular disease, chronic kidney disease, chronic obstructive pulmonary disease, dementia, diabetes, dyslipidemia, hypertension, an immunocompromised status, a mental health disorder, obesity, prior SARS-CoV-2 infection, early vaccination status, and pandemic wave (measured on the day follow-up commenced). The proportional hazards assumption of the models was assessed using the Supremum test and Schoenfeld residuals, and violations were addressed as necessary. To account for potential event dependence due to multiple outcomes within individuals, Marginal Cox regression with the robust sandwich covariance matrix was evaluated. As results were similar to those assuming independent events, standard Cox regression models were retained for simplicity. For sub-group analysis of those who received ≥1 additional dose, time-varying additional dose and pandemic wave status were incorporated in the Cox models.

Examples of characteristics that may be used to define at-risk populations or ‘risk profiles’ were evaluated among the primary vaccination schedule cohort. The number of individuals who had a severe COVID-19 outcome, the proportion of all severe COVID-19 outcome events that occurred among the cohort for which individuals in the risk profile represent, population size, and uHR (95% CI) for each risk profile was determined. Population attributable risk was calculated based on observed incidence and risk profile prevalence to estimate the proportion of severe COVID-19 outcomes potentially preventable.

In accordance with data custodian privacy standards, outcomes with 1 to 9 individuals where there was a potential for identification were reported as <10; where applicable, other outcomes were censored (e.g., presented as a range; associated proportions were presented based on the mid number of the range) so the number of individuals within the small cell size outcome could not be calculated and potentially identified [33]. A 2-sided significance level of 0.05 was applied for all statistical tests. Analysis was performed using R (version 4.4.0) statistical software [34].

## 3. Results

### 3.1. Cohort selection

Among 3,639,134 adults who had ≥1 COVID-19 vaccination dose during the inclusion period, the cohort consisted of 2,639,731 adults who had a primary 2 dose vaccination schedule (or 1 dose of Ad26.COV2.S); the sub-group contained 1,580,110 adults who had ≥1 additional dose (Figure 1; Supplementary Figure 1 shows data linkage). The majority of the cohort completed their primary vaccination schedule during the period of Alpha variant predominance (57.0%; February 15, 2021 to July 14, 2021), followed by Delta (37.8%; July 15, 2021 to November 27, 2021); 78.8% of those who received their first additional dose did so during the period of Omicron variant predominance (November 28, 2021 onwards; Table 1).

**Figure 1.**
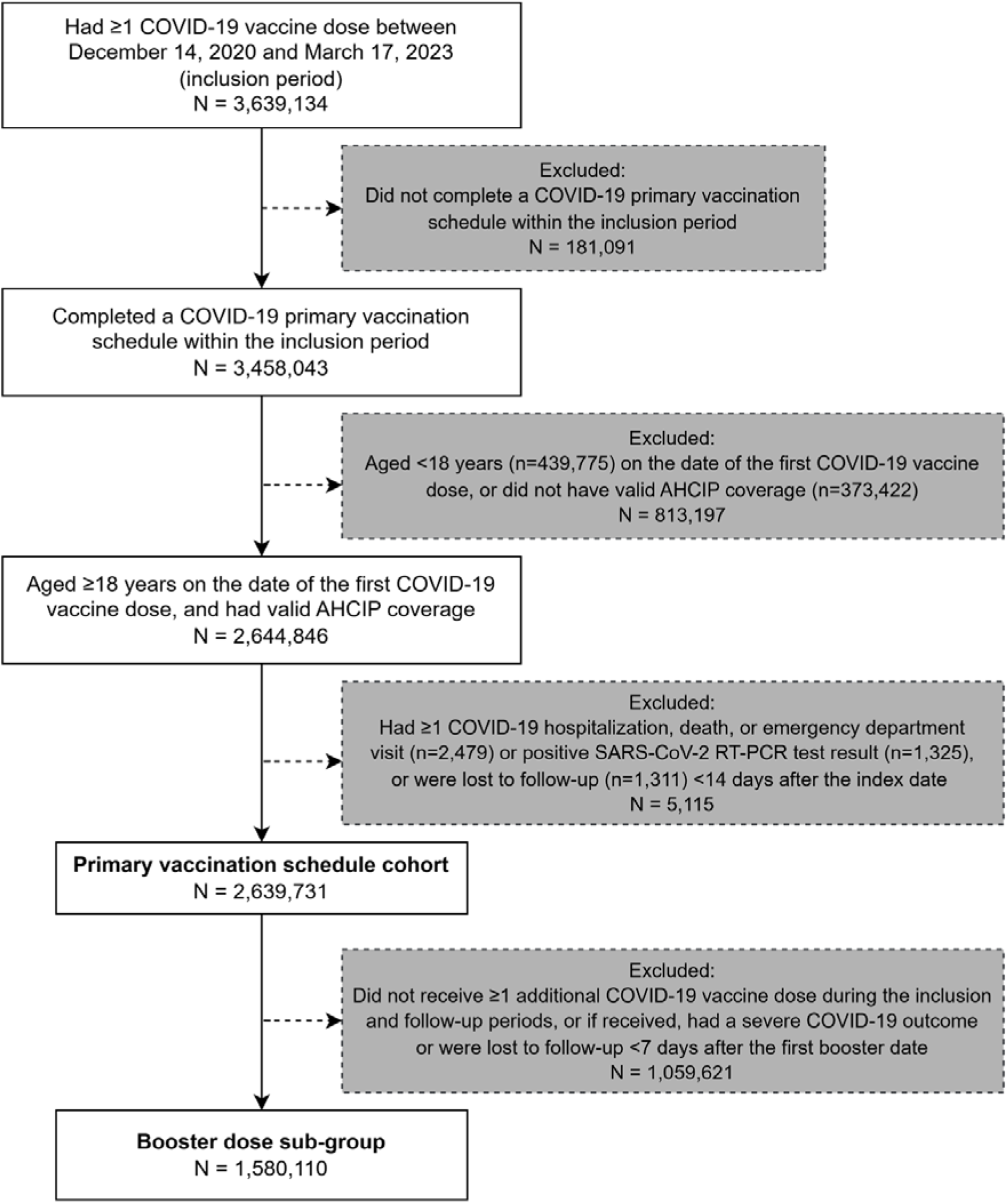
Cohort selection flow diagram. Abbreviations: AHCIP = Alberta Health Care Insurance Plan; COVID-19 = coronavirus disease 2019; RT-PCR = reverse transcription polymerase chain reaction; SARS-CoV-2 = severe acute respiratory syndrome coronavirus 2.

**Table 1.** Baseline characteristics.

| | Cohort<br>Received a primary<br>vaccination schedule<br>n=2,639,731 | Sub-group<br>Received $\geq 1$ additional<br>dose<br>n=1,580,110 |
| --- | --- | --- |
| **Demographic characteristics** |  |  |
| Age (years) |  |  |
| Overall |  |  |
| Mean (SD) | 49 (18) | 54 (18) |
| Median (Q1-Q3) | 48 (34-62) | 55 (40-67) |
| Category, n (%) |  |  |
| 18-39 | 922,177 (34.9%) | 383,021 (24.2%) |
| 40-49 | 480,430 (18.2%) | 256,288 (16.2%) |
| 50-59 | 455,363 (17.3%) | 296,755 (18.8%) |
| 60-64 | 227,907 (8.6%) | 172,813 (10.9%) |
| $\geq 65$ | 553,854 (21.0%) | 471,233 (29.8%) |
| Sex, n (%) |  |  |
| Female | 1,344,497 (50.9%) | 843,652 (53.4%) |
| Male | ~1,295,229 (49.1%) | ~736,453 (46.6%) |
| Other | <10 (0.0%) | <10 (0.0%) |
| Residence, n (%) |  |  |
| Rural/urban |  |  |
| Rural | 310,696 (11.8%) | 159,525 (10.1%) |
| Urban | 2,329,035 (88.2%) | 1,420,585 (89.9%) |
| Long-term care | 13,711 (0.5%) | 10,999 (0.7%) |
| **Socioeconomic status, n (%)** |  |  |
| Material deprivation index |  |  |
| 1 (most well off) | 505,914 (19.2%) | 342,788 (21.7%) |
| 2 | 499,729 (18.9%) | 313,791 (19.9%) |
| 3 | 496,310 (18.8%) | 294,177 (18.6%) |
| 4 | 515,065 (19.5%) | 294,648 (18.6%) |
| 5 (most deprived) | 495,583 (18.8%) | 255,849 (16.2%) |
| Missing | 127,130 (4.8%) | 78,857 (5.0%) |
| **Clinical characteristics** |  |  |
| Charlson Comorbidity Index |  |  |
| Score |  |  |
| Mean (SD) | 0.5 (1.2) | 0.7 (1.3) |
| Median (Q1-Q3) | 0 (0-1) | 0 (0-1) |
| Category; comorbid burden, n (%) |  |  |
| 0; none | 1,907,314 (72.3%) | 1,050,473 (66.5%) |
| 1-2; mild | 582,467 (22.1%) | 411,414 (26.0%) |
| 3-4; moderate | 102,610 (3.9%) | 82,059 (5.2%) |
| $\geq 5$; severe | 47,340 (1.8%) | 36,164 (2.3%) |
| Health conditions of interest, n (%) |  |  |
| Dyslipidemia | 646,915 (24.5%) | 457,263 (28.9%) |
| Hypertension | 612,715 (23.2%) | 468,007 (29.6%) |
| Mental health disorder | 408,786 (15.5%) | 242,562 (15.4%) |
| Cardiovascular disease | 277,654 (10.5%) | 194,388 (12.3%) |
| Diabetes | 274,180 (10.4%) | 204,686 (13.0%) |
| Immunocompromised (overall) | 268,507 (10.2%) | 184,778 (11.7%) |
| Autoimmune disorders | 89,294 (3.4%) | 63,142 (4.0%) |
| Cancer | 41,659 (1.6%) | 32,964 (2.1%) |
| Dialysis | 2,967 (0.1%) | 2,413 (0.2%) |
| History of a transplant | 10,985 (0.4%) | 8,747 (0.6%) |
| Others | 215,618 (8.2%) | 148,240 (9.4%) |
| Obesity | 228,273 (8.6%) | 148,728 (9.4%) |
| Chronic kidney disease | 146,502 (5.5%) | 114,864 (7.3%) |
| COPD | 127,072 (4.8%) | 94,767 (6.0%) |
| Asthma | 99,895 (3.8%) | 65,176 (4.1%) |
| Dementia | 40,880 (1.5%) | 33,214 (2.1%) |
| Parkinson's disease | 7,709 (0.3%) | 6,337 (0.4%) |
| Pregnancy | 6,235 (0.2%) | 2,990 (0.2%) |
| Pulmonary hypertension | 2,304 (0.1%) | 1,714 (0.1%) |
| Down's syndrome | 1,083 (0.0%) | 802 (0.1%) |
| **Other factors, n (%)** |  |  |
| Prior SARS-CoV-2 infection | 143,337 (5.4%) | 59,051 (3.7%) |
| Received nirmatrelvir/ritonavir | 20,336 (0.8%) | 18,161 (1.1%) |
| Early vaccination | 2,454,057 (93.0%) | 1,572,946 (99.5%) |
| Pandemic wave vaccine dose received* |  |  |
| Wave 1 | 0 (0.0%) | 0 (0.0%) |
| Wave 2 | 15,447 (0.6%) | 0 (0.0%) |
| Wave 3 (Alpha) | 1,504,004 (57.0%) | 856 (0.1%) |
| Wave 4 (Delta) | 998,956 (37.8%) | 334,221 (21.2%) |
| Wave 5 (Omicron) | 121,324 (4.6%) | 1,245,033 (78.8%) |

\*Pandemic wave was measured on the day follow-up commenced. Wave 1 = March 1, 2020 to June 30, 2020; Wave 2 = July 1, 2020 to February 14, 2021; Wave 3 = February 15, 2021 to July 14, 2021; Wave 4 = July 15, 2021 to November 27, 2021; Wave 5 = November 28, 2021 onwards. Abbreviations: COPD = chronic obstructive pulmonary disease; SARS-CoV-2 = severe acute respiratory syndrome coronavirus 2; SD = standard deviation; Q = quintile.

On the index date, the mean age of the cohort was 49 (SD 18) years, and 50.9% were female; the most common (>10%) health conditions of interest that individuals were living with were dyslipidemia (24.5%), hypertension (23.2%), a mental health disorder (15.5%), cardiovascular disease (10.5%), diabetes (10.4%), and an overall immunocompromised status (included all immunocompromising conditions; 10.2%) (Table 1). Mean age of the sub-group was 54 (SD 18) years, and 53.4% were female (Table 1).

### 3.2. Primary vaccination schedule cohort

Overall, 5,475 adults (0.2% of the cohort; cumulative 1-year incidence: 28.5 per 10,000 persons; incidence density rate: 2.2 events per 1,000 person-years [Supplementary Table 4]) had ≥1 severe COVID-19 outcome, including 970 who died (0.04% of the cohort; 5.3 per 10,000 persons; 0.4 deaths per 1,000 person-years [Supplementary Table 4]). Supplementary Table 5 details the expected number of individuals who had a severe COVID-19 outcome at 6-, 12-, and 24-months after completion of a primary vaccination schedule.

In unadjusted analysis (Figure 2), the likelihood of a severe COVID-19 outcome was greater in those who were older (uHR [95% CI]; versus 18-39 years: 2.09 [1.88-2.32] in those 50-59 years, 3.82 [3.40-4.29] in those 60-64 years, 12.51 [11.56-13.56] in those ≥65 years; ≥65 versus <65 years: 8.83 [8.33-9.35]), male (versus female: 1.07 [1.01-1.13]), resided in a rural area (versus urban: 1.56 [1.46-1.68]), long-term care facility (versus not: 8.75 [7.25-10.57]), or a socioeconomically deprived area (most deprived versus most well-off: 1.78 [1.63-1.94]), and had a higher comorbid burden (versus one score lower: 1.51 [1.50-1.53]) or were living with a health condition of interest (ranged from 1.33 [1.25-1.42] to 29.25 [23.14-36.99]; see Figure 2 for results of each condition) The likelihood of a severe COVID-19 outcome was lower in those who had a prior SARS-CoV-2 infection (versus not; 0.56 [0.49-0.65]) (Figure 2). Supplementary Table 6 shows results by type of severe COVID-19 outcome (hospitalization [non-intensive care unit], intensive care unit admission, death).

**Figure 2.**
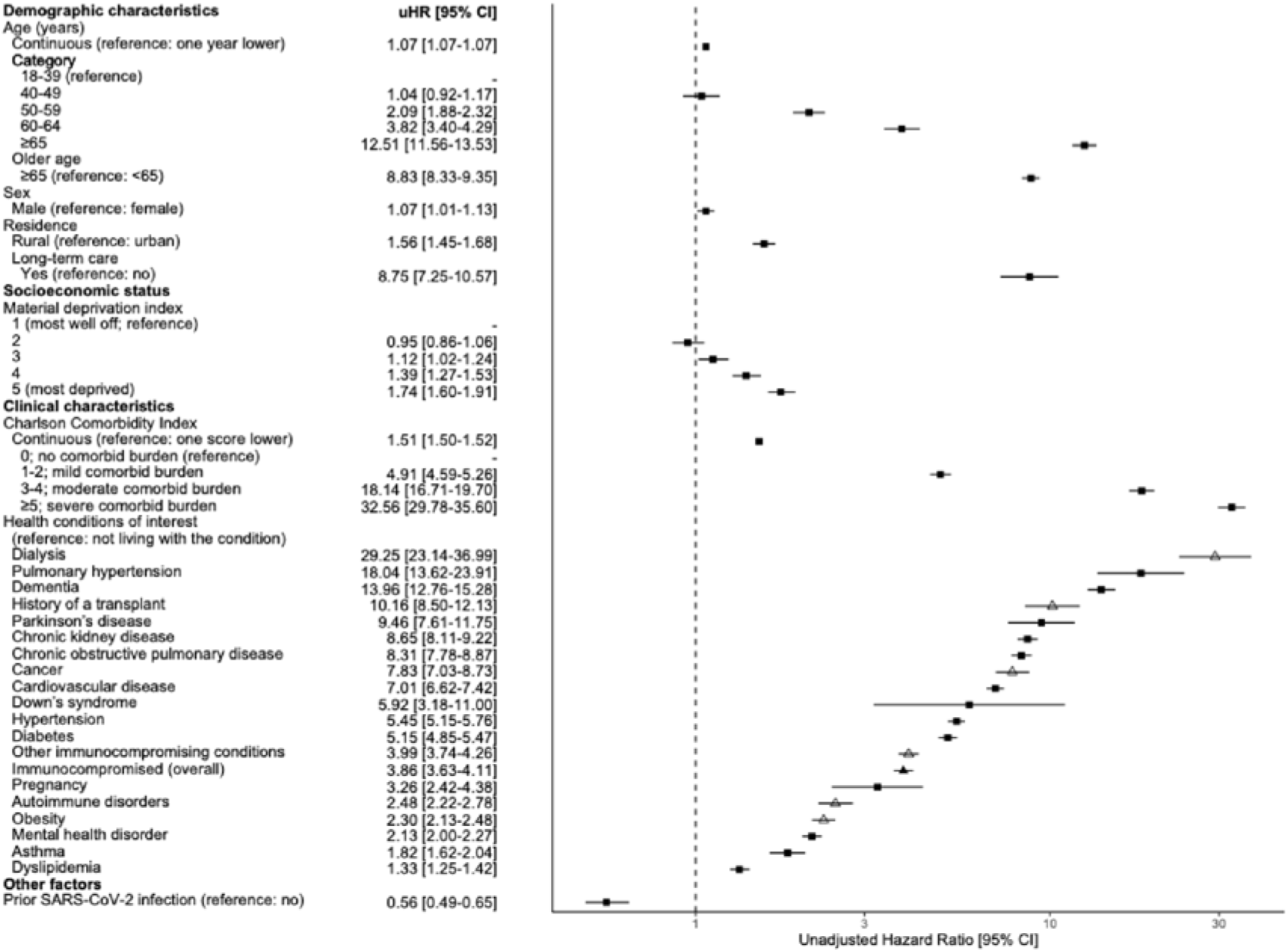
Unadjusted likelihood of a severe COVID-19 outcome (any type) among adults who completed a primary vaccination series. Triangle symbols show immunocompromised conditions. Abbreviations: CI = confidence interval; COVID-19 = coronavirus disease 2019; HR = hazard ratio; SARS-CoV-2 = severe acute respiratory syndrome coronavirus 2

In fully adjusted analysis (adjusted for sociodemographic characteristics, overall comorbid burden, select health conditions of interest, prior SARS-CoV-2 infection, early vaccination status, and pandemic wave; Table 2), the likelihood of a severe COVID-19 outcome was greater in those who were ≥65 versus <65 years (aHR [95% CI]; 3.56 [3.30-3.84]), male (versus female: 1.14 [1.08-1.20]), resided in a long-term care facility (versus not for a COVID-19 related death: 2.03 [1.55-2.66]) or a socioeconomically deprived area (most well-off versus most deprived: 1.39 [1.27-1.52]), and had a higher comorbid burden (versus once score lower: 1.14 [1.12-1.16]) or were living with a health condition of interest (versus not) (included 10 conditions; ranged from 1.17 [1.08-1.27] in those living with obesity to 2.07 [1.87-2.30] in those living with dementia); dyslipidemia was negatively associated (0.77 [0.72-0.82]). Rural residence (versus urban: 1.05 [0.98-1.13]) was not significant (Table 2). The likelihood of a severe COVID-19 outcome was lower in those who had a prior SARS-CoV-2 infection (versus not: 0.62 [0.54-0.72]) (Table 2).

**Table 2.**
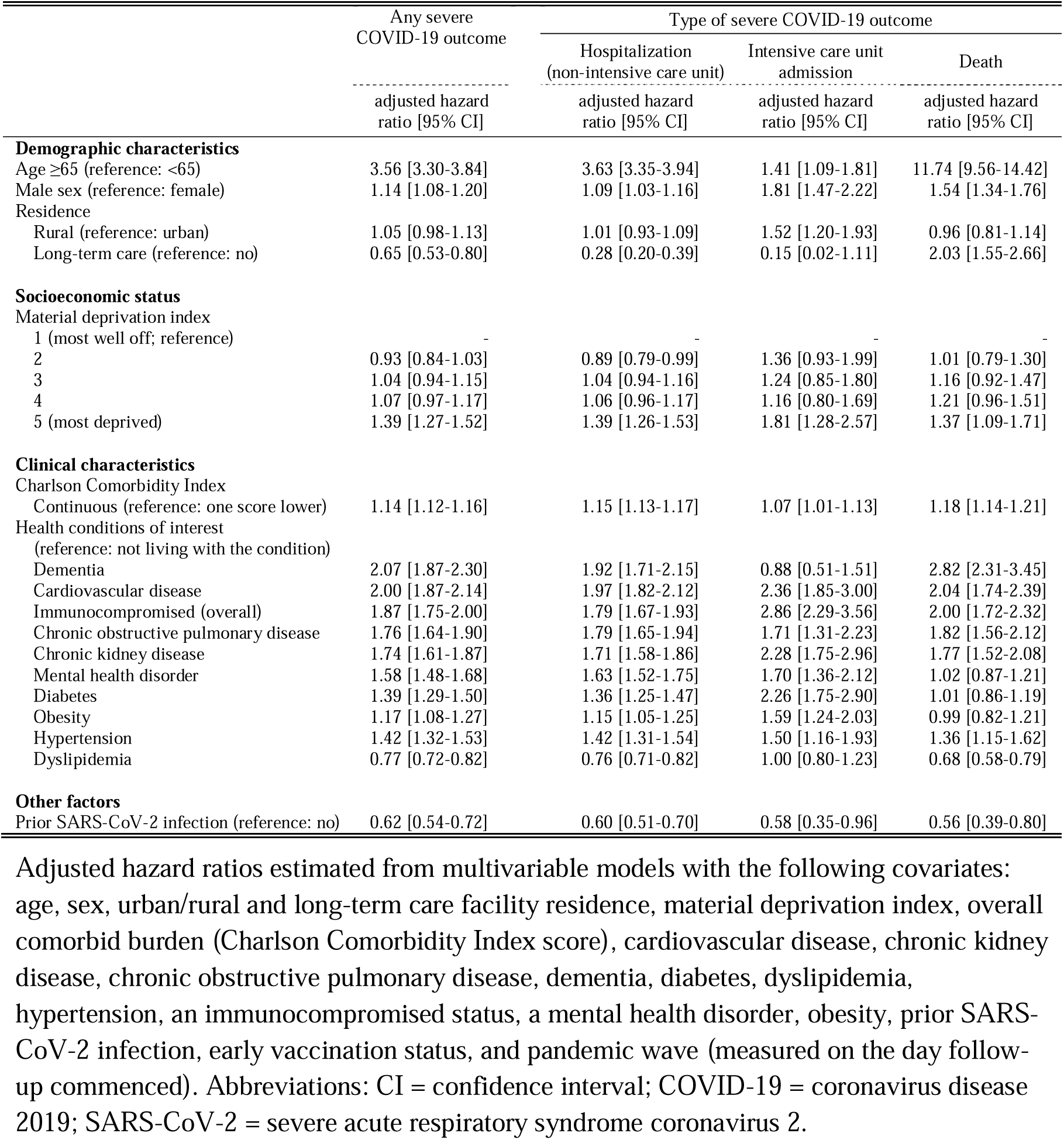
Adjusted likelihood of a severe COVID-19 outcome among adults who completed a primary vaccination series.

|  | Any severe<br>COVID-19 outcome | Type of severe COVID-19 outcome |  |  |
| --- | --- | --- | --- | --- |
|  |  | Hospitalization<br>(non-intensive care unit) | Intensive care unit<br>admission | Death |
|  | adjusted hazard<br>ratio [95% CI] | adjusted hazard<br>ratio [95% CI] | adjusted hazard<br>ratio [95% CI] | adjusted hazard<br>ratio [95% CI] |
| **Demographic characteristics** |  |  |  |  |
| Age ≥65 (reference: <65) | 3.56 [3.30-3.84] | 3.63 [3.35-3.94] | 1.41 [1.09-1.81] | 11.74 [9.56-14.42] |
| Male sex (reference: female) | 1.14 [1.08-1.20] | 1.09 [1.03-1.16] | 1.81 [1.47-2.22] | 1.54 [1.34-1.76] |
| Residence |  |  |  |  |
| Rural (reference: urban) | 1.05 [0.98-1.13] | 1.01 [0.93-1.09] | 1.52 [1.20-1.93] | 0.96 [0.81-1.14] |
| Long-term care (reference: no) | 0.65 [0.53-0.80] | 0.28 [0.20-0.39] | 0.15 [0.02-1.11] | 2.03 [1.55-2.66] |
| **Socioeconomic status** |  |  |  |  |
| Material deprivation index |  |  |  |  |
| 1 (most well off; reference) | - | - | - | - |
| 2 | 0.93 [0.84-1.03] | 0.89 [0.79-0.99] | 1.36 [0.93-1.99] | 1.01 [0.79-1.30] |
| 3 | 1.04 [0.94-1.15] | 1.04 [0.94-1.16] | 1.24 [0.85-1.80] | 1.16 [0.92-1.47] |
| 4 | 1.07 [0.97-1.17] | 1.06 [0.96-1.17] | 1.16 [0.80-1.69] | 1.21 [0.96-1.51] |
| 5 (most deprived) | 1.39 [1.27-1.52] | 1.39 [1.26-1.53] | 1.81 [1.28-2.57] | 1.37 [1.09-1.71] |
| **Clinical characteristics** |  |  |  |  |
| Charlson Comorbidity Index |  |  |  |  |
| Continuous (reference: one score lower) | 1.14 [1.12-1.16] | 1.15 [1.13-1.17] | 1.07 [1.01-1.13] | 1.18 [1.14-1.21] |
| Health conditions of interest<br>(reference: not living with the condition) |  |  |  |  |
| Dementia | 2.07 [1.87-2.30] | 1.92 [1.71-2.15] | 0.88 [0.51-1.51] | 2.82 [2.31-3.45] |
| Cardiovascular disease | 2.00 [1.87-2.14] | 1.97 [1.82-2.12] | 2.36 [1.85-3.00] | 2.04 [1.74-2.39] |
| Immunocompromised (overall) | 1.87 [1.75-2.00] | 1.79 [1.67-1.93] | 2.86 [2.29-3.56] | 2.00 [1.72-2.32] |
| Chronic obstructive pulmonary disease | 1.76 [1.64-1.90] | 1.79 [1.65-1.94] | 1.71 [1.31-2.23] | 1.82 [1.56-2.12] |
| Chronic kidney disease | 1.74 [1.61-1.87] | 1.71 [1.58-1.86] | 2.28 [1.75-2.96] | 1.77 [1.52-2.08] |
| Mental health disorder | 1.58 [1.48-1.68] | 1.63 [1.52-1.75] | 1.70 [1.36-2.12] | 1.02 [0.87-1.21] |
| Diabetes | 1.39 [1.29-1.50] | 1.36 [1.25-1.47] | 2.26 [1.75-2.90] | 1.01 [0.86-1.19] |
| Obesity | 1.17 [1.08-1.27] | 1.15 [1.05-1.25] | 1.59 [1.24-2.03] | 0.99 [0.82-1.21] |
| Hypertension | 1.42 [1.32-1.53] | 1.42 [1.31-1.54] | 1.50 [1.16-1.93] | 1.36 [1.15-1.62] |
| Dyslipidemia | 0.77 [0.72-0.82] | 0.76 [0.71-0.82] | 1.00 [0.80-1.23] | 0.68 [0.58-0.79] |
| **Other factors** |  |  |  |  |
| Prior SARS-CoV-2 infection (reference: no) | 0.62 [0.54-0.72] | 0.60 [0.51-0.70] | 0.58 [0.35-0.96] | 0.56 [0.39-0.80] |

In the example risk profiles considered (Figure 3), the majority of severe COVID-19 outcomes occurred among those aged ≥65 years (54.7%), who represented 21.0% of adults who received a primary vaccination schedule. The largest proportion of severe COVID-19 outcomes occurred among those aged ≥65 years or living with any health condition of interest (regardless of age) (89.6%), representing 54.7% of adults who received a primary vaccination schedule, the largest population attributable risk (77.0%), and a relatively high likelihood of a severe COVID-19 outcome (uHR [95% CI]; 9.34 [8.57-10.19]) (Figure 3). Supplementary Table 7 shows results by type of severe COVID-19 outcome (hospitalization [non-intensive care unit], intensive care unit admission, death).

**Figure 3.**
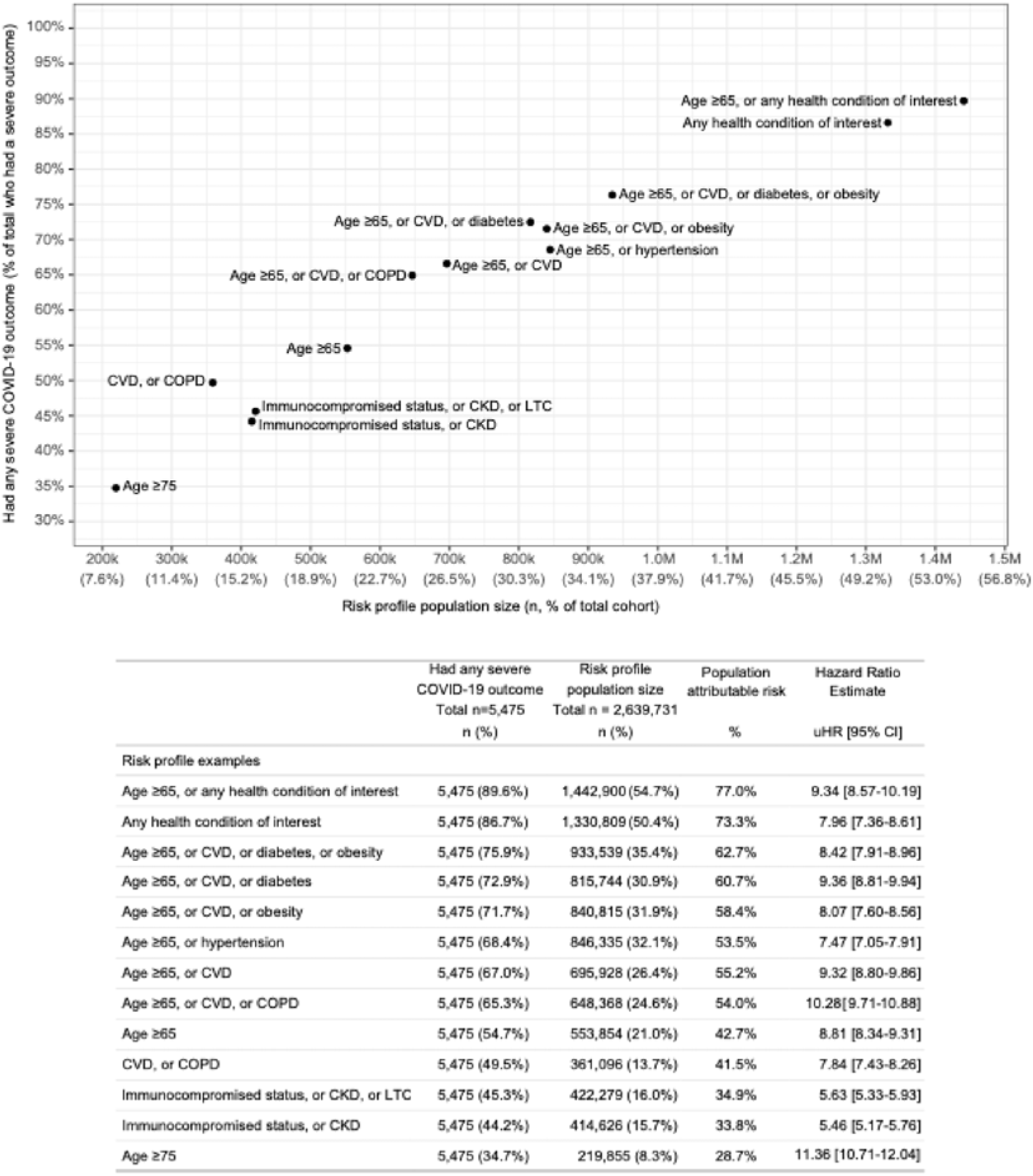
Examples of unadjusted risk profiles among adults who completed a primary vaccination series. Any health condition of interest included asthma, cardiovascular disease, chronic kidney disease, chronic obstructive pulmonary disease, dementia, diabetes, Down’s syndrome, hypertension, an immunocompromised status, a mental health disorder, obesity, Parkinson’s disease, pregnancy, and pulmonary hypertension; dyslipidemia was not included. Hazard ratio estimates are unadjusted, with the reference group comprising individuals without the specified condition(s). Population attributable risk is unadjusted and based on observed incidence and risk factor prevalence in the cohort, estimating the proportion of severe COVID-19 outcomes potentially preventable by targeting these factors. Abbreviations: CI = confidence interval; CKD = chronic kidney disease; COPD = chronic obstructive pulmonary disease; COVID-19 = coronavirus disease 2019; CVD = cardiovascular disease; LTC = long-term care; uHR = unadjusted hazard ratio.

### 3.3. Additional dose sub-group

In unadjusted analysis (Table 3), the likelihood of a severe COVID-19 outcome among the sub-group who received ≥1 additional dose was similar to unadjusted results of the primary vaccination schedule cohort, with the exception of those living with dyslipidemia which was not a significant risk (uHR [95% CI]: 1.04 [0.99-1.09]). In fully adjusted analysis (Table 4), the likelihood of a severe COVID-19 outcome was similar to adjusted results of the primary vaccination schedule cohort, with the exception of socioeconomic status (most well-off versus most deprived: aHR [95% CI]: 1.04 [0.97-1.11]) and living with obesity (1.01 [0.94-1.09]) – these factors were not significant.

**Table 3.** Unadjusted likelihood of a severe COVID-19 outcome among adults who received ≥1 additional dose.

|  | Any severe COVID-19 outcome | Type of severe COVID-19 outcome |  |  |
| --- | --- | --- | --- | --- |
|  |  | Hospitalization (non-intensive care unit) | Intensive care unit admission | Death |
|  | adjusted hazard ratio [95% CI] | adjusted hazard ratio [95% CI] | adjusted hazard ratio [95% CI] | adjusted hazard ratio [95% CI] |
| **Demographic characteristics** |  |  |  |  |
| Age (years) |  |  |  |  |
| Continuous (reference: one year lower) | 1.10 [1.10-1.10] | 1.10 [1.10-1.10] | 1.06 [1.05-1.06] | 1.15 [1.14-1.15] |
| Category |  |  |  |  |
| 18-39 (reference) | - | - | - | - |
| 40-49 | 1.24 [1.06-1.45] | 1.09 [0.91-1.29] | 3.34 [1.94-5.76] | 3.09 [1.59-6.02] |
| 50-59 | 2.60 [2.28-2.97] | 2.30 [1.99-2.65] | 6.10 [3.69-10.08] | 8.26 [4.57-14.91] |
| 60-64 | 4.71 [4.10-5.40] | 4.18 [3.61-4.83] | 10.23 [6.13-17.09] | 19.07 [10.62-34.25] |
| $\geq 65$ | 26.44 [23.69-29.52] | 23.62 [21.07-26.48] | 19.73 [12.34-31.53] | 189.77 [109.90-327.70] |
| Older age |  |  |  |  |
| $\geq 65$ (reference: <65) | 13.67 [12.95-14.44] | 13.48 [12.71-14.31] | 4.82 [3.99-5.84] | 33.86 [29.11-39.39] |
| Sex |  |  |  |  |
| Male (reference: female) | 1.31 [1.25-1.37] | 1.33 [1.27-1.39] | 1.77 [1.47-2.13] | 1.28 [1.17-1.40] |
| Residence |  |  |  |  |
| Rural (reference: urban) | 1.59 [1.50-1.69] | 1.54 [1.44-1.65] | 2.07 [1.64-2.61] | 1.64 [1.45-1.85] |
| Long-term care (reference: no) | 13.16 [11.92-14.53] | 5.86 [5.02-6.83] | 4.21 [2.08-8.53] | 48.32 [42.50-54.93] |
| **Socioeconomic status** |  |  |  |  |
| Material deprivation index |  |  |  |  |
| 1 (most well off; reference) | - | - | - | - |
| 2 | 0.93 [0.84-1.03] | 0.89 [0.79-0.99] | 1.36 [0.93-1.99] | 1.01 [0.79-1.30] |
| 3 | 1.04 [0.94-1.15] | 1.04 [0.94-1.16] | 1.24 [0.85-1.80] | 1.16 [0.92-1.47] |
| 4 | 1.07 [0.97-1.17] | 1.06 [0.96-1.17] | 1.16 [0.80-1.69] | 1.21 [0.96-1.51] |
| 5 (most deprived) | 1.39 [1.27-1.52] | 1.39 [1.26-1.53] | 1.81 [1.28-2.57] | 1.37 [1.09-1.71] |
| **Clinical characteristics** |  |  |  |  |
| Charlson Comorbidity Index |  |  |  |  |
| Continuous (reference: one score lower) | 1.14 [1.12-1.16] | 1.15 [1.13-1.17] | 1.07 [1.01-1.13] | 1.18 [1.14-1.21] |
| Category; comorbid burden, n (%) |  |  |  |  |
| 0; none (reference) | - | - | - | - |
| 1-2; mild | 5.79 [5.45-6.15] | 5.37 [5.03-5.74] | 5.11 [4.00-6.52] | 10.18 [8.78-11.81] |
| 3-4; moderate | 20.56 [19.22-22.00] | 19.36 [17.99-20.82] | 16.68 [12.60-22.08] | 36.29 [30.96-42.54] |
| $\geq 5$; severe | 38.36 [35.70-41.21] | 35.93 [33.24-38.84] | 37.25 [28.04-49.49] | 76.77 [65.30-90.24] |
| Health conditions of interest (reference: not living with the condition) |  |  |  |  |
| Dialysis | 24.79 [21.67-28.36] | 25.64 [22.18-29.63] | 44.73 [29.38-68.08] | 16.16 [11.71-22.28] |
| Dementia | 14.75 [13.87-15.68] | 11.08 [10.29-11.93] | 3.58 [2.30-5.57] | 39.23 [35.51-43.33] |
| Pulmonary hypertension | 14.66 [11.96-17.97] | 15.04 [12.06-18.76] | 18.92 [8.96-39.94] | 12.06 [7.76-18.75] |
| History of transplant | 11.66 [10.51-12.94] | 11.70 [10.44-13.11] | 29.89 [22.30-40.07] | 9.98 [8.00-12.44] |
| Parkinson's disease | 10.81 [9.55-12.23] | 10.41 [9.06-11.96] | 2.16 [0.69-6.73] | 13.42 [10.75-16.75] |
| Chronic kidney disease | 7.65 [7.29-8.03] | 7.79 [7.39-8.21] | 7.03 [5.73-8.62] | 7.96 [7.22-8.77] |
| Cardiovascular disease | 7.54 [7.21-7.88] | 7.43 [7.08-7.80] | 7.79 [6.48-9.36] | 8.83 [8.07-9.66] |
| Chronic obstructive pulmonary disease | 6.69 [6.36-7.04] | 6.86 [6.49-7.25] | 5.71 [4.57-7.13] | 6.62 [5.97-7.34] |
| Cancer | 6.15 [5.70-6.64] | 6.34 [5.83-6.89] | 6.94 [5.11-9.43] | 6.82 [5.88-7.90] |
| Hypertension | 5.70 [5.44-5.97] | 5.73 [5.44-6.03] | 4.72 [3.90-5.70] | 6.70 [6.07-7.40] |
| Other immunocompromising conditions | 4.00 [3.80-4.20] | 4.04 [3.83-4.27] | 6.58 [5.45-7.94] | 3.83 [3.46-4.24] |
| Immunocompromised (overall) | 3.98 [3.79-4.17] | 3.99 [3.79-4.20] | 6.78 [5.64-8.15] | 3.83 [3.48-4.21] |
| Diabetes | 3.74 [3.57-3.92] | 3.88 [3.69-4.08] | 4.40 [3.64-5.33] | 3.34 [3.03-3.68] |
| Down's syndrome | 2.68 [1.44-4.98] | 2.57 [1.28-5.13] | 9.11 [2.27-36.52] | 1.10 [0.16-7.84] |
| Autoimmune disorders | 2.40 [2.22-2.60] | 2.42 [2.22-2.64] | 3.08 [2.28-4.17] | 2.14 [1.80-2.53] |
| Mental health disorder | 1.82 [1.73-1.92] | 1.77 [1.68-1.88] | 1.74 [1.40-2.15] | 1.81 [1.63-2.01] |
| Asthma | 1.62 [1.47-1.77] | 1.69 [1.54-1.87] | 1.90 [1.33-2.70] | 0.92 [0.73-1.17] |
| Obesity | 1.51 [1.40-1.62] | 1.55 [1.43-1.67] | 2.02 [1.54-2.64] | 1.13 [0.96-1.34] |
| Dyslipidemia | 1.03 [0.99-1.09] | 1.07 [1.02-1.13] | 1.21 [0.99-1.47] | 0.79 [0.71-0.88] |
| Pregnancy | 0.66 [0.36-1.24] | 0.80 [0.43-1.48] | NA* | NA* |
| **Other factors** |  |  |  |  |
| Prior SARS-CoV-2 infection (reference: no) | 0.62 [0.54-0.72] | 0.60 [0.51-0.70] | 0.58 [0.35-0.96] | 0.56 [0.39-0.80] |

\*The number of events were not sufficient to calculate the hazard ratio. Abbreviations: CI = confidence interval; COVID-19 = coronavirus disease 2019; NA = not applicable; SARS-CoV-2 = severe acute respiratory syndrome coronavirus 2.

**Table 4.** Adjusted likelihood of a severe COVID-19 outcome among adults who received ≥1 additional dose.

|  | Any severe<br>COVID-19 outcome | Type of severe COVID-19 outcome |  |  |
| --- | --- | --- | --- | --- |
|  |  | Hospitalization<br>(non-intensive care unit) | Intensive care unit<br>admission | Death |
|  | adjusted hazard<br>ratio [95% CI] | adjusted hazard<br>ratio [95% CI] | adjusted hazard<br>ratio [95% CI] | adjusted hazard<br>ratio [95% CI] |
| **Demographic characteristics** |  |  |  |  |
| Age $\geq 65$ (reference: $<65$) | 5.01 [4.70-5.34] | 5.08 [4.74-5.44] | 1.56 [1.25-1.95] | 11.00 [9.32-12.99] |
| Male sex (reference: female) | 1.25 [1.19-1.31] | 1.24 [1.18-1.31] | 1.65 [1.36-1.99] | 1.29 [1.18-1.42] |
| Residence |  |  |  |  |
| Rural (reference: urban) | 0.91 [0.86-0.97] | 0.89 [0.83-0.95] | 1.20 [0.94-1.53] | 0.89 [0.78-1.01] |
| Long-term care (reference: no) | 1.11 [1.00-1.24] | 0.54 [0.46-0.64] | 0.95 [0.44-2.05] | 2.48 [2.14-2.87] |
| **Socioeconomic status** |  |  |  |  |
| Material deprivation index |  |  |  |  |
| 1 (most well off; reference) | - | - | - | - |
| 2 | 0.98 [0.91-1.05] | 0.97 [0.89-1.05] | 0.81 [0.58-1.12] | 1.08 [0.93-1.25] |
| 3 | 1.01 [0.94-1.08] | 1.00 [0.93-1.08] | 1.03 [0.76-1.40] | 1.05 [0.91-1.22] |
| 4 | 1.06 [0.99-1.13] | 1.05 [0.98-1.13] | 1.16 [0.87-1.55] | 1.11 [0.96-1.27] |
| 5 (most deprived) | 1.04 [0.97-1.11] | 1.01 [0.94-1.09] | 1.38 [1.05-1.82] | 1.08 [0.94-1.25] |
| **Clinical characteristics** |  |  |  |  |
| Charlson Comorbidity Index |  |  |  |  |
| Continuous (reference: one score lower) | 1.12 [1.11-1.14] | 1.12 [1.11-1.14] | 1.15 [1.10-1.20] | 1.15 [1.13-1.18] |
| Health conditions of interest<br>(reference: not living with the condition) |  |  |  |  |
| Dementia | 2.34 [2.18-2.51] | 2.02 [1.86-2.19] | 0.77 [0.47-1.26] | 4.24 [3.74-4.80] |
| Cardiovascular disease | 1.95 [1.86-2.06] | 1.96 [1.86-2.08] | 2.30 [1.85-2.86] | 1.81 [1.63-2.01] |
| Immunocompromised (overall) | 1.93 [1.84-2.03] | 1.90 [1.80-2.00] | 3.39 [2.78-4.14] | 1.88 [1.70-2.08] |
| Chronic obstructive pulmonary disease | 1.62 [1.54-1.71] | 1.66 [1.57-1.76] | 1.69 [1.34-2.13] | 1.42 [1.28-1.58] |
| Chronic kidney disease | 1.57 [1.48-1.65] | 1.64 [1.55-1.74] | 1.44 [1.13-1.83] | 1.30 [1.17-1.45] |
| Mental health disorder | 1.41 [1.33-1.49] | 1.41 [1.33-1.50] | 1.52 [1.21-1.91] | 1.34 [1.20-1.49] |
| Diabetes | 1.37 [1.30-1.44] | 1.40 [1.32-1.48] | 1.41 [1.13-1.76] | 1.11 [0.99-1.24] |
| Obesity | 1.12 [1.06-1.18] | 1.17 [1.11-1.25] | 1.39 [1.11-1.74] | 0.85 [0.76-0.95] |
| Hypertension | 1.01 [0.94-1.09] | 1.02 [0.94-1.10] | 1.14 [0.86-1.50] | 0.84 [0.71-1.00] |
| Dyslipidemia | 0.75 [0.72-0.79] | 0.76 [0.72-0.80] | 0.85 [0.69-1.03] | 0.64 [0.57-0.71] |
| **Other factors** |  |  |  |  |
| Prior SARS-CoV-2 infection (reference: no) | 0.46 [0.39-0.54] | 0.51 [0.43-0.61] | 0.19 [0.07-0.51] | 0.29 [0.20-0.43] |

Adjusted hazard ratios estimated from multivariable models with the following covariates: age, sex, urban/rural and long-term care facility residence, material deprivation index, overall comorbid burden (Charlson Comorbidity Index score), cardiovascular disease, chronic kidney disease, chronic obstructive pulmonary disease, dementia, diabetes, dyslipidemia, hypertension, an immunocompromised status, a mental health disorder, obesity, prior SARS-CoV-2 infection, early vaccination status, and time-varying additional dose and associated pandemic wave (measured on the day follow-up commenced). Abbreviations: CI = confidence interval; COVID-19 = coronavirus disease 2019; SARS-CoV-2 = severe acute respiratory syndrome coronavirus 2.

## 4. Discussion

In this real world retrospective, observational, population-based cohort study, results showed that among adults who received a primary vaccination schedule between December 2020 and March 2023 in Alberta, Canada, severe COVID-19 outcomes were low (0.2% of the cohort). Factors that were associated with a relatively greater risk of a severe COVID-19 outcome among the primary vaccination schedule cohort included older age (≥50 years; particularly ≥65 years), and living with a higher comorbid burden or health condition of interest. Among the health conditions examined, most displayed a similar magnitude of risk across the measurement approaches. Those with the greatest magnitude of risk (top 10 unadjusted associations listed for completeness) included receiving dialysis, living with pulmonary hypertension, dementia, a history of solid organ or bone marrow transplant, Parkinson’s disease, chronic kidney disease, chronic obstructive pulmonary disease, cancer and receiving chemotherapy or an immunocompromising drug, cardiovascular disease, and Down’s syndrome. Other factors that were associated with risk of a severe COVID-19 outcome included male sex, and residing in a rural area (unadjusted analysis only; suggests that the distribution of other factors, and not residence location, is mediating the effect), long-term care facility (COVID-19 related death in adjusted analysis), or socioeconomically deprived area. Factors associated with risk among those who received ≥1 additional dose were largely similar to those identified in the primary vaccination schedule cohort. As the definition of ‘high risk’ can vary depending on the context, results from the different analytical approaches used in this study provide complementary insights into factors that may be associated with a severe COVID-19 outcome (unadjusted analysis) which can be used for individual or clinical decision making, along with examples of the relative contribution of risk groups to the overall burden of severe outcomes (population attributable risk) which can be used as a template for public health planning; how factors may exert their effects is informed by fully adjusted analyses.

Previous studies have been conducted to identify risk factors for developing a severe COVID-19 outcome among those who have received a primary vaccination schedule or additional doses. Older age consistently appears to be strongly and increasingly associated with risk of a severe COVID-19 outcome [12–19]. For example, Semenzato et al., (2022) found that among 28 million individuals in France who received a primary vaccination schedule by July, 2021, risk rose steadily in those aged 55-64 years and older (reference: 45-54 years), such that those aged 85-89 years had a 4-fold (aHR 4.02 [3.47-4.65]) increased risk of hospitalization and 38-fold (aHR 37.95 [19.15-75.23]) increased risk of death [13]. Among the factors evaluated in this study, older age (≥50 years; particularly ≥65 years) was found to be strongly associated with a relatively greater risk of a severe COVID-19 outcome, in unadjusted and fully adjusted analyses, among those who received at least a primary vaccination schedule. Considering that, in this study, adults aged ≥65 years accounted for the majority (54.7%) of those who had a severe COVID-19 outcome, and comprised 21.0% of the cohort population, this age group assumed a disproportionately high burden of severe outcomes. Collectively, these findings further reinforce the importance of prioritizing older adults in infection prevention strategies and public health campaigns, irrespective of comorbidity burden or underlying health condition. Findings also provide empirical support for Canada’s current National Advisory Committee on Immunisation (NACI) recommendations for guidance on the use of COVID-19 vaccines, which advise that *all* adults aged ≥65 years remain up to date with COVID-19 vaccination to mitigate the risk of severe outcomes.

Individuals who have received at least a primary vaccination schedule and are living with a higher comorbid burden or certain health conditions have been shown to be at higher risk of a severe COVID-19 outcome [12–19]. In these studies, most of the health conditions examined were found to be associated with increased risk; among the health conditions, dialysis, immunosuppressive therapy, a history of transplant, neurological disease, several active cancers, chronic respiratory disease, chronic kidney disease, and cardiovascular disease were consistently reported to be most strongly associated with an increased risk of a severe COVID-19 outcome among those who received a primary vaccination schedule [12–15], and ≥1 additional dose [17–19]. Similar findings were observed in this study, and support current NACI recommendations that advise up to date COVID-19 vaccination for individuals with underlying health conditions that place them at higher risk of a severe COVID-19 outcome [4]. Of note, while individuals living with an overall immunocompromised status in this study were found to be at risk of a severe COVID-19 outcome, risk markedly varied among sub-groups. For example, among the primary vaccination schedule cohort, risk ranged from 2-times greater in those living with an autoimmune disorder such as arthritis or inflammatory bowel disease (uHR: 2.48 [2.22-2.78]) to 29-times greater in those receiving dialysis (uHR: 29.25 [23.14-36.99]), which is consistent with other reports [17, 35]. A more detailed understanding of the risk of a severe COVID-19 outcome associated with various types and degrees of immunocompromise – whether related to cancer, immunosuppressive therapies, or diseases directly affecting the immune system – enables optimal communication tailored to the public, specific patient groups, and health care providers. Similarly, improved knowledge of the relative impact of common and less common specific health conditions in the population informs both accurate individual risk stratification and facilitates refined vaccination and prevention strategies at the population level.

A number of risk profiles that may be considered for health and public health interventions using alternate age and health condition criteria were explored in this study. Identifying target populations for public health interventions such as vaccination involves trade-offs between capturing the majority of those at elevated risk, the feasibility of deploying the intervention (complexity of recommendation and extent of targeting less common but high risk conditions), and size of population implicated with attendant resource implications. Decision maker understanding of these trade-offs may allow transparent optimization of recommendations given resource constraints.

This study has several important strengths including the large size and population-based design. However, results need to be interpreted in the context of the limitations of administrative data sets. Retrospective claims-based studies use administrative data (e.g., vaccination records, codes, laboratory results) as opposed to medical records (contain detailed notes including diagnoses), and therefore, there is a potential for misclassification of study cohorts or measures; validated case definitions were applied to administrative data, where available, to address this limitation. Individuals meeting medical criteria for obesity were likely underrepresented in this study, as codes for obesity have been reported to have low sensitivity, but high specificity and positive predictive value. [36]. Those who had a prior SARS-CoV-2 infection, which would impact their immune response and severity of subsequent infection were also likely underrepresented, as administrative health data does not include community-based rapid antigen test results. Although ethnicity has been reported as a factor in severe COVID-19 outcomes among those vaccinated [14], this variable is not included in Alberta administrative health data, and therefore could not be assessed. Finally, this study focused on adults with at least a primary vaccination schedule between December 14, 2020 and March 17, 2023, and therefore is not generalizable to those <18 years, those who are unvaccinated or under vaccinated, and those who received further vaccine doses or acquired newer COVID-19 variants after the study period.

## 5. Conclusions

In a population protected by a primary vaccination schedule, results from this real world study showed that although severe COVID-19 outcomes were not frequent, an elevated risk of a severe COVID-19 outcome remained in those who were older (≥50 years; particularly ≥65 years), and those who were living with a higher comorbid burden or health condition of interest in Alberta, Canada. Other factors that were associated with risk included male sex, and residing in a rural area (unadjusted analysis), long-term care facility, or socioeconomically deprived area. Results were largely similar among those who received ≥1 additional dose. These uniquely Canadian findings support current recommendations from NACI regarding COVID-19 vaccination; specifically, they reinforce guidance that advise all individuals at increased risk of a severe COVID-19 outcome – such as adults ≥65 years, those with certain health conditions, residents of long-term care and other congregated living settings, and members of equity-denied communities – should remain up to date with their COVID-19 vaccinations to reduce the risk of a severe COVID-19 outcome [4]. Among the risk profile examples, alternate groupings of characteristics resulted in trade-offs between the size of sub-population identified and the fraction of population with a severe COVID-19 outcome captured.

This study illustrates how evidence from real world population-level health system data can be used to guide individual and clinical decision making, inform public health communication and tailored messaging, and support policy development aimed at protecting those who remain at elevated risk of severe COVID-19 outcomes in a largely vaccinated population. Even after vaccination, individuals with identifiable risk factors may be prioritized for ongoing preventative strategies; these may include timely administration of additional vaccination doses, the management of underlying health conditions, mitigation of exposure risk, provision of COVID-19 treatments when appropriate, and conducting educational, community, and health care professional outreach initiatives. Continued updated assessment and surveillance of factors that may be associated with the absolute and relative risk of a severe COVID-19 outcome is recommended, analogous to approaches used for other endemic illnesses such as influenza.

## Declarations

### Ethics approval

Ethics approval was received from the University of Alberta Research Ethics Board (Pro00135542) that granted an exemption from requiring written informed consent (a waiver of consent was applied).

### Consent to participate

A waiver of consent was applied by the University of Alberta Research Ethics Board (Pro00135542).

### Consent for publication

Not applicable.

### Data availability statement

The data that support the findings of this study are available from AHS and Alberta Health, but restrictions apply to the availability of these data, which were used under license for the current study, and so are not publicly available.

### Code availability

Analysis was performed using R (version 4.4.0).

### Author Contributions Statement

All authors contributed to the study concept. HL drafted the protocol, and performed quality control analytics. SAH led data acquisition, conducted the analyses, and created the tables and figures. KM prepared the draft manuscript. JR provided project management. All authors contributed to the interpretation of the data and critical revision of the manuscript for intellectual content; academic and clinical researchers retained the right to accept or reject comments or suggestions from funders. SK provided study supervision.

## Acknowledgements

We thank the participants of this study. This study is based in part on data from Alberta Health and AHS, which was provided by the Alberta Strategy for Patient Oriented Research (SPOR) SUPPORT Unit. The interpretation and conclusions contained herein are those of the authors and do not necessarily represent the views or opinions of the Government of Alberta or AHS. SK was supported by the Kidney Health Research Chair and the Division of Nephrology at the University of Alberta.

