## Supplementary for "Factors associated with severe COVID-19 outcomes among adults with at least a primary vaccination schedule: a retrospective cohort study from Alberta, Canada"

Supplementary material table of contents

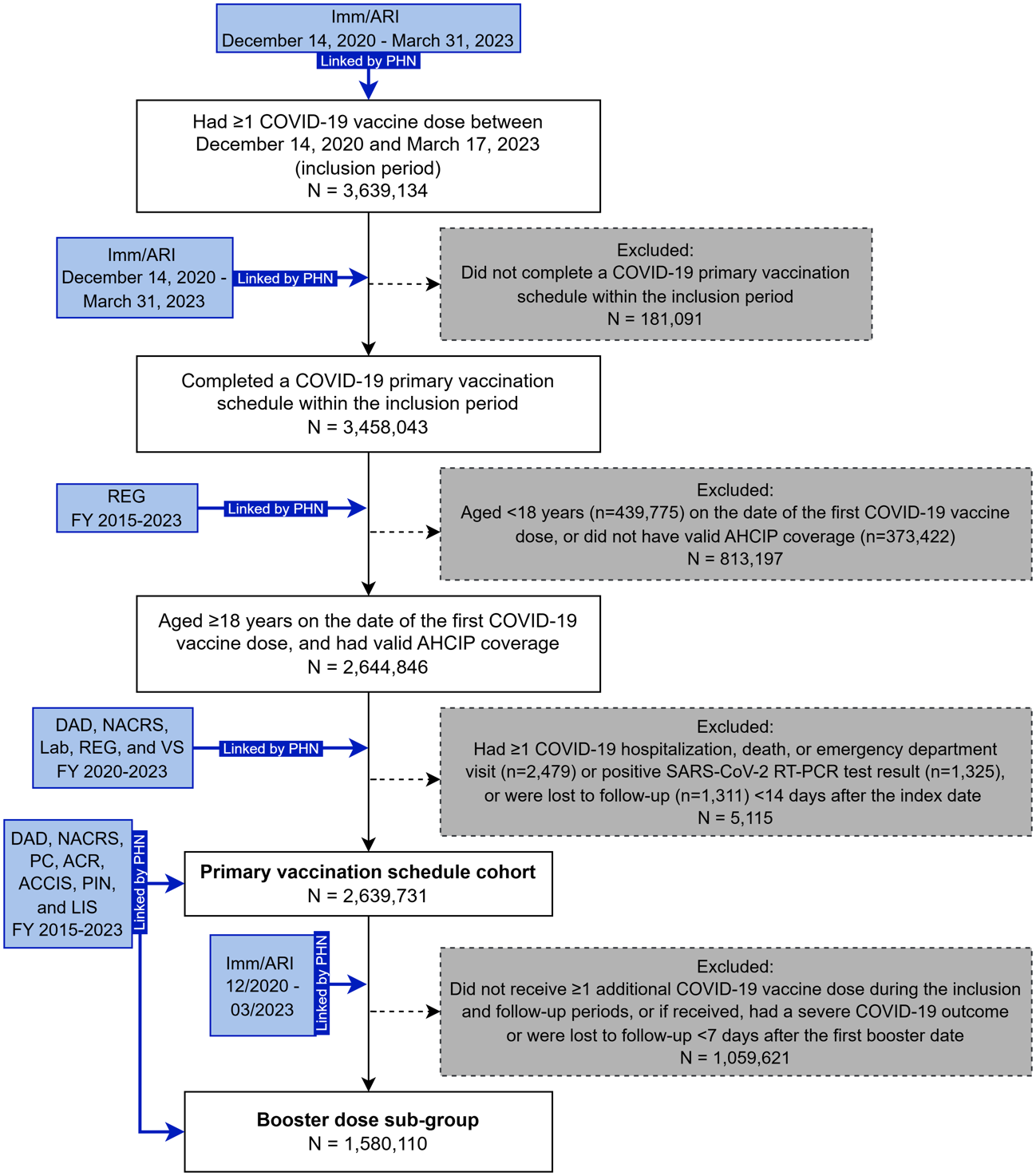

Supplementary Figure 1. Cohort selection flow diagram with associated data linkage. Abbreviations: ACCIS = Alberta Continuing Care Information System; AHCIP = Alberta Health Care Insurance Plan; ACR = Alberta Cancer Registry; COVID-19 = coronavirus disease 2019; DAD = Discharge Abstract Database; FY = fiscal year; Imm/ARI = Immunization and Adverse Reactions to Immunizations; LIS = Laboratory Information System; NACRS = National Ambulatory Care Reporting System; PC = Practitioner Claims; PHN = Personal Health Number; PIN = Pharmaceutical Information Network; REG = Population Registry; RT-PCR = reverse transcription polymerase chain reaction; SARS-CoV-2 = severe acute respiratory syndrome coronavirus 2; VS = Vital Statistics.

### Supplementary Table 1. Health conditions and their associated codes and weights included in the Charlson Comorbidity Index.

| Health condition | ICD-9-CM codes | ICD-10-CA codes | Weight |
| --- | --- | --- | --- |
| Myocardial infarction | 410, 412 | I21, I22, I25.2 | 1 |
| Congestive heart failure | 398, 402, 425, 428 | I09.9, I11.0, I13.0, I13.2, I25.5, I42.0, I42.5, I42.6, I42.7, I42.8, I42.9, I43, I50, P29.0 | 1 |
| Peripheral vascular disease | 440, 441, 443, 447, 557 | I70, I71, I73.1, I73.8, I73.9, I77.1, I79.0, I79.2, K55.1, K55.8, K55.9, Z95.8, Z95.9 | 1 |
| Cerebrovascular disease | 430, 431, 432, 433, 434, 435, 436, 437, 438 | G45, G46, I60, I61, I62, I63, I64, I65, I66, I67, I68, I69, H34.0 | 1 |
| Dementia | 290, 294, 331 | F00, F01, F02, F03, G30, F05.1, G31.1 | 1 |
| Chronic pulmonary disease | 416, 490, 491, 492, 493, 494, 495, 496, 500, 501, 502, 503, 504, 505 | J40, J41, J42, J43, J44, J45, J46, J47, J60, J61, J62, J63, J64, J65, J66, J67, I27.8, I27.9, J68.4, J70.1, J70.3 | 1 |
| Connective tissue disease | 446, 710, 714, 725 | M05, M32, M33, M34, M06, M31.5, M35.1, M35.3, M36.0 | 1 |
| Peptic ulcer disease | 531, 532, 533, 534 | K25, K26, K27, K28 | 1 |
| Mild liver disease | 070, 570, 571, 573 | B18, K73, K74, K70.0, K70.1, K70.2, K70.3, K70.9, K71.7, K71.3, K71.4, K71.5, K76.0, K76.2, K76.3, K76.4, K76.8, K76.9, Z94.4 | 1 |
| Moderate/severe liver disease | 456, 572 | K70.4, K71.1, K72.1, K72.9, K76.5, K76.6, K76.7, I85.0, I85.9, I86.4, I98.2 | 3 |
| Diabetes (without complication) | 250 | E10.0, E10.l, E10.6, E10.8, E10.9, E11.0, E11.1, E11.6, E11.8, E11.9, E12.0, E12.1, E12.6, E12.8, E12.9, E13.0, E13.1, E13.6, E13.8, E13.9, E14.0, E14.1, E14.6, E14.8, E14.9 | 1 |
| Diabetes (with complication) | 250 | E10.2, E10.3, E10.4, E10.5, E10.7, E11.2, E11.3, E11.4, E11.5, E11.7, E12.2, E12.3, E12.4, E12.5, E12.7, E13.2, E13.3, E13.4, E13.5, E13.7, E14.2, E14.3, E14.4, E14.5, E14.7 | 2 |
| Hemiplegia and paraplegia | 334, 342, 343, 344 | G81, G82, G04.1, G11.4, G80.1, G80.2, G83.0, G83.1, G83.2, G83.3, G83.4, G83.9 | 2 |
| Moderate or severe renal disease | 403, 582, 583, 585, 586, 588, V56 | N18, N19, N05.2, N05.3, N05.4, N05.5, N05.6, N05.7, N25.0, I12.0, I13.1, N03.2, N03.3, N03.4, N03.5, N03.6, N03.7, Z49.0, Z49.1, Z49.2, Z94.0, Z99.2 | 2 |
| Cancer | 140-165, 170-172, 174-176, 179-195, 200- 208, 238 | C00-C26, C30-C34, C37-C41, C43, C45-C58, C60-C76, C81-C85, C88, C90- C97 | 2 |
| Metastatic Carcinoma | 196, 197, 198, 199 | C77, C78, C79, C80 | 6 |
| HIV/AIDS | 042, 043, 044 | B20, B21, B22, B24 | 6 |

To be considered as having one of the listed diseases, a participant must have ≥1 hospitalization (associated ICD-10-CA code listed in any diagnostic field) or ≥2 physician claims (associated ICD-9-CM codes listed in any diagnostic field) of the corresponding ICD within ≤2-years. Abbreviations: HIV/AIDS = Human immunodeficiency virus/acquired immunodeficiency syndrome; ICD-9-CM = International classification of diseases, ninth revision, clinical modification; ICD-10-CA = International classification of diseases, tenth revision, Canadian enhancement

### Supplementary Table 2. Administrative data case definitions used to identify health conditions of interest.

| Health condition | Algorithm | ICD-9-CM codes | ICD-10-CA codes; other codes |
| --- | --- | --- | --- |
| Asthma | ≥1 hospitalization or ≥3 ambulatory care / claims in ≤2 years | 493 | J45 |
| Cardiovascular disease (any of the below) | |  |  |
| Atrial fibrillation | ≥1 hospitalization or ≥2 claims in ≤2 years | 427.3 | I48.0 |
| Chronic heart failure | ≥1 hospitalization or ≥2 claims in ≤2 years | 398.9, 402, 404, 425.4 –425.9, 428 | I09.9, I25.5, I42.0, I42.5–I42.9, I43, I50 |
| Coronary artery disease | ≥1 hospitalization or ≥1 ambulatory care visit or ≥1 procedure or ≥2 claims in ≤2 years | 410-413 | I20–I25; *Procedure codes:* 1.IJ.57.GQ, 1.IJ.50, 1.IL.35, 1.IJ.76 |
| Peripheral artery disease | ≥1 hospitalization or ≥1 ambulatory care visit or ≥1 claim in any years | 440.2 | I70.2 |
| Stroke | ≥1 hospitalization or emergency department visit with a stroke code in the most responsible diagnostic field OR ≥1 hospitalization, emergency department visit, or ambulatory care visit* with a stroke code in a secondary diagnostic field and a z-code in the most responsible field in any years. |  | G08, G45 (excluding subcode G45.4), H34.0, H34.1, I60, I61, I62.9, I63, I64, I67.6; Z50 (excluding subcodes Z50.2, Z50.3, Z50.4), *Z51.5 (only applies to ambulatory care), Z54.8, Z54.9 |
| Chronic kidney disease | ≥1 hospitalization or ≥3 claims in ≤1 year OR mean eGFR <60 mL/min*1.73 m^2^ or mean albuminuria >30 mg/g over 1-year | 583-586, 592, 593.9 | N00-N23; *Laboratory results:* eGFR and albuminuria |
| Chronic obstructive pulmonary disease | ≥35 years with ≥1 hospitalization or claim in any years | 491-492, 496 | J41–J44 |
| Dementia | ≥1 hospitalization or ≥2 claims in ≤2 years | 290, 294.1, 331.2 | F00–F03, F05.1, G30, G31.1 |
| Diabetes | ≥1 hospitalization or ≥2 claims in ≤2 years | 250 | E10-E14 |
| Down’s syndrome | ≥1 hospitalization, ambulatory visit, or claim in any year | 758 | Q90 |
| Dyslipidemia | ≥1 hospitalization, ambulatory care visit, claim, or lab in ≤3 years; condition present for 3 years | 272 | E78; *Laboratory* *results*: LDL ≥3.5mmol/L or non-HDL ≥4.3mmol/L or apolipoprotein B100 ≥1.2 g/L |
| Hypertension | ≥1 hospitalization or ≥2 claims in ≤2 years | 401-405 | I10-I13, I15 |
| Immunocompromised (any of the below) | | | |
| Autoimmune disorders |  |  |  |
| Inflammatory bowel disease | ≥2 hospitalizations, ≥2 ambulatory care visits, or ≥4 claims in ≤2 years | 555, 556 | K50, K51 |
| Multiple sclerosis | ≥1 hospitalization, or ≥5 claims from ambulatory care / claims in ≤2 years | 340 | G35 |
| Psoriasis | ≥1 hospitalization or ≥2 claims | 696 | L40.0-L40.4, L40.8, L40.9 |
| Psoriatic arthritis | ≥1 hospitalization or ≥3 claims for seronegative SpA (≥1 by a rheumatologist, internal medicine) and ≥1 claim for psoriasis | 696 (psoriasis), 721 (SpA) | L40.5, M07.0-M07.3, M09.0 |
| Rheumatoid arthritis | ≥1 hospitalization or ≥3 claims with ≥1 by a specialist (rheumatologist, orthopedic surgeon, internal medicine) in ≤2 years | 714 | M05, M06 |
| Systematic autoimmune rheumatoid disease | ≥1 hospitalization or ≥3 claims | 710 | M32-M34, M35.0, M35.8, M35.9, M36.0 |
| Cancer and receiving chemotherapy or an immunocompromising drug | ≥1 hospitalization or ≥2 claims in ≤2 years and received chemotherapy or an immunocompromising drug ≤1 year before index date | 140-165, 170-172, 174-176, 179-195, 196-199, 200-208, 238 | C00-C26, C30-C34, C37-C41, C43, C45-C58, C60-C76, C77-C80, C81-C85, C88, C90-C97, D01.0-D01.3, D02.2, D05-D06, D07.5; *Health services code for chemotherapy:* 13.55A; *Procedure codes for chemotherapy:* 1.ZZ.35.CA-M0, 1.ZZ.35.CA-M5, 1.ZZ.35.CA-M9, 1.ZZ.35.HA-M0, 1.ZZ.35.HA-M5, 1.ZZ.35.HA-M9, 1.ZZ.35.YA-M0, 1.ZZ.35.YA-M5, 1.ZZ.35.YA-M9; *Drug codes:* see bottom of table |
| History of solid organ or bone marrow transplant | ≥1 procedure | V42.0-V42.2, V42.4, V42.6-V42.9 | Z48.2, Z94 (not including: Z94.5, Z94.83, Z94.88, Z94.9); *Procedure codes:* 1.PC.85, 1.OA.85, 1.HY.85, 1.HZ.85, 1.GR.85, 1.OJ.85, 1.GT.85, 1.OK.85, 1.NK.85, 1.NP.85, 1.WY.19, 1.LZ.19.HH-U7, 1.LZ.19.HH-U8 |
| Receiving dialysis | ≥1 hospitalization or ambulatory care visit in ≤2 years before the index date | V45.1, V56 | Z49, Z99.2; *Health services code:* 13.99 A, B, C, D, O, or OA |
| Other immunocompromising conditions | |  |  |
| Diseases of the blood | ≥1 hospitalization |  | D70-D72, D73.0-D73.2 |
| Human immunodeficiency virus | ≥1 hospitalization or ≥3 claims in ≤3 years | 042-044 | B20-B24 |
| Immune system disorders | ≥1 hospitalization, emergency department visit , or claim OR ≥1 dispensation for an immunocompromising drug ≤1 year before index date | 279 | D80-D84, D89; *Drug codes:* see bottom of table |
| Sickle cell anemia | ≥1 hospitalization or ≥2 claims in ≤1 year | 282.6 | D57.0-D57.2, D57.8 |
| Spleen malformations or removal | ≥1 hospitalization |  | Q89.0; *Procedure codes:* 1.OB.87, 1.OB.89 |
| Treated for tuberculosis | ≥1 hospitalization or ≥1 claim | 010-018 | A15-A19 |
| A mental health disorder (any of the below) | |  |  |
| Bipolar disorder | ≥1 hospitalization OR ≥3 claims OR ≥1 claim and ≥3 dispensations for bipolar disorder treatment drugs in ≤5 years | 296 | F31; *Drug codes:* ATC N05AN01, N03AF01, N03AG01, N03AX09 |
| Depression | ≥1 hospitalization or ≥2 claims in ≤2 years | 300.4, 311 | F32, F33, F34.1 |
| Schizophrenia | ≥1 hospitalization or ≥2 claims in ≤2 years | 295 | F20, F21, F23.2, F25 |
| Obesity | ≥1 hospitalization, ambulatory visit, or claim in any years (ICD or BMI fee-modifier code in any years); excluded if no code listed after most recent dispensation for a weight-loss medication OR had bariatric surgery | 278.0, V77.8 | E66; *BMI modifier code:* BMI^^^; *Drug codes:* ATC A08AB01, A08AA62; or DIN 02437899; *Health services codes:* 55.8A, 55.8B 55.9A, or 56.93; *Procedure code:* 1.NF.78.^^ |
| Parkinson’s disease | ≥40 years with ≥1 hospitalization; OR ≥2 claims (at least 30 days between claims); OR ≥1 hospitalization / claim and ≥ 1 drug dispensation for Parkinson’s disease treatment in ≤1 year | 332 | G20, G21.0, G21.1, G21.2, G21.3, G21.4, G21.8, G21.9, G22, F02.3; *Drug codes:* ATC N04BD01, N04BD01, N04BA01, N04BA03 |
| Pregnant | Female sex with ≥1 hospitalization, ambulatory visit, or claim with ICD code for pregnancy OR delivery ≤6 months after index date (excluding abortion / miscarriage ICD codes) | 632-637, 640-648, 650-665, V22, V23, V27 | O00, O02.1, O10-O16, O20-O29, O30-O48, O60-O75, Z34, Z35, Z37; *Health services codes:* 87.0X, 87.2X; *Procedure codes:* 5.CA.88, 5.CA.89, 5.CA.90, 5.CA.93, 5.MD.5, 5.MD.60 |
| Pulmonary hypertension | ≥1 hospitalization | 416 | I27 |
| ATC codes for immunocompromising drugs: H02AA02, H02AB01, H02AB02, H02AB04, H02AB06, H02AB07, H02AB08, H02AB09, H02AB10, H02AB57, H02BX01, L01AA01, L01AA02, L01AA03, L01AA05, L01AA06, L01AA09, L01AB01, L01AB02, L01AC01, L01AD01, L01AD02, L01AD04, L01AX03, L01AX04, L01BA01, L01BA03, L01BA04, L01BA05, L01BB02, L01BB03, L01BB04, L01BB05, L01BB06, L01BB07, L01BC01, L01BC02, L01BC05, L01BC06, L01BC07, L01BC08, L01BC52, L01BC59, L01CA01, L01CA02, L01CA03, L01CA04, L01CB01, L01CB02, L01CD01, L01CD02, L01CD04, L01CE01, L01CE02, L01CX01, L01DA01, L01DB01, L01DB02, L01DB03, L01DB06, L01DB07, L01DB09, L01DC01, L01DC03, L01EA01, L01EA02, L01EA03, L01EA04, L01EA05, L01EA06, L01EB01, L01EB02, L01EB03, L01EB04, L01EB07, L01EC01, L01EC02, L01EC03, L01ED01, L01ED02, L01ED03, L01ED04, L01ED05, L01EE01, L01EE02, L01EE03, L01EE04, L01EF01, L01EF02, L01EF03, L01EG01, L01EG02, L01EH01, L01EH02, L01EH03, L01EJ01, L01EJ02, L01EK01, L01EL01, L01EL02, L01EL03, L01EM01, L01EM03, L01EN01, L01EN02, L01EN03, L01EX01, L01EX02, L01EX03, L01EX04, L01EX05, L01EX07, L01EX08, L01EX09, L01EX10, L01EX12, L01EX13, L01EX14, L01EX17, L01EX19, L01EX21, L01EX22, L01EX23, L01FA, L01FA01, L01FA02, L01FA03, L01FB01, L01FC01, L01FC02, L01FD01, L01FD02, L01FD03, L01FD04, L01FE01, L01FE02, L01FE03, L01FF01, L01FF02, L01FF03, L01FF04, L01FF05, L01FF06, L01FF07, L01FG01, L01FG02, L01FX02, L01FX03, L01FX04, L01FX05, L01FX06, L01FX07, L01FX08, L01FX09, L01FX10, L01FX12, L01FX13, L01FX14, L01FX17, L01FX18, L01X, L01XA01, L01XA02, L01XA03, L01XB01, L01XD01, L01XD03, L01XD04, L01XE, L01XF01, L01XG01, L01XG02, L01XG03, L01XH01, L01XH02, L01XJ01, L01XJ03, L01XK01, L01XK02, L01XK04, L01XL03, L01XL04, L01XL05, L01XL06, L01XL07, L01XX01, L01XX02, L01XX03, L01XX05, L01XX08, L01XX11, L01XX23, L01XX24, L01XX27, L01XX35, L01XX41, L01XX44, L01XX52, L01XX59, L01XX66, L01XX69, L01XX73, L01XX74, L01XX75, L01XY01, L01XY02, L02BA01, L02BA03, L02BB01, L02BB02, L02BB03, L02BB04, L02BB05, L02BB06, L02BG01, L02BG02, L02BG03, L02BG04, L02BG06, L02BX02, L02BX03, L03AA02, L03AA12, L03AA13, L03AB, L03AC01, L03AX03, L03AX12, L03AX13, L03AX16, L04AA02, L04AA03, L04AA04, L04AA06, L04AA10, L04AA13, L04AA15, L04AA18, L04AA21, L04AA23, L04AA24, L04AA25, L04AA26, L04AA27, L04AA29, L04AA31, L04AA32, L04AA33, L04AA34, L04AA36, L04AA37, L04AA38, L04AA40, L04AA42, L04AA43, L04AA44, L04AA47, L04AA48, L04AA50, L04AA51, L04AA52, L04AA54, L04AA56, L04AA59, L04AB01, L04AB02, L04AB04, L04AB05, L04AB06, L04AC01, L04AC02, L04AC03, L04AC05, L04AC07, L04AC08, L04AC10, L04AC11, L04AC12, L04AC13, L04AC14, L04AC16, L04AC17, L04AC18, L04AC19, L04AC21, L04AC22, L04AD01, L04AD02, L04AX01, L04AX02, L04AX03, L04AX04, L04AX06, V10XX02, X10XA53. | | | |

Case definitions were applied over a 5-year period before the index date, unless otherwise stated. Abbreviations: ATC = anatomical therapeutic chemical; BMI = body mass index; DIN = drug identification number; eGFR = estimated glomerular filtration rate; HDL = high-density lipoprotein; ICD-9-CM: International Classification of Disease – Version 9 – Clinical Modification (Alberta specific); ICD-10-CA: International Classification of Disease – Version 10 – Canadian Enhancement; LDL = low-density lipoprotein; SpA = spondyloarthropathies.

### Supplementary Table 3. ICD-10-CA codes used to define a COVID-19-related hospital admission.

| ICD-10-CA code | Description |
| --- | --- |
| *Code located within any diagnostic field:* | |
| U07.1 | COVID-19 virus identified. Assigned for COVID-19 that is confirmed by a positive COVID-19 test result, or when the physician or primary care provider or infection control staff documented a positive COVID-19 test result. |
| *Code located in a secondary diagnostic field:* | |
| U07.3 | Multisystem inflammatory syndrome associated with COVID-19. Assigned when the patient is diagnosed with multisystem inflammatory syndrome associated with COVID-19. |
| *Code located in the most responsible diagnostic field and a positive SARS-CoV-2 RT-PCR test result ≤14 days prior to or during the hospital admission:* | |
| J04 | Acute laryngitis and tracheitis |
| J09 | Influenza due to identified Avian influenza virus |
| J10 | Influenza due to other identified influenza virus |
| J11 | Influenza, virus not identified |
| J12 | Viral pneumonia, not elsewhere classified |
| J13 | Pneumonia due to Streptococcus pneumoniae |
| J14 | Pneumonia due to Haemophilus influenzae |
| J15 | Bacterial pneumonia, not elsewhere classified |
| J16 | Pneumonia due to other infectious organisms, not elsewhere classified |
| J17 | Pneumonia in diseases classified elsewhere |
| J18 | Pneumonia, organism unspecified |
| J20 | Acute bronchitis |
| J21 | Acute bronchiolitis |
| J22 | Unspecified acute lower respiratory infection |
| J80 | Adult respiratory distress syndrome |
| U04 | Severe acute respiratory syndrome [SARS] |

Abbreviations: COVID-19 = coronavirus disease 2019; ICD-10-CA = International Classification of Disease – Version 10 – Canadian Enhancement ; RT-PCR = reverse transcription polymerase chain reaction; SARS-CoV-2 = severe acute respiratory syndrome coronavirus 2.

### Supplementary Table 4. Cumulative 1-year incidence and incidence density rates of severe COVID-19 outcome events among adults who completed a primary vaccination series.

|  | All severe COVID-19 outcomes | | | | |  | Type of severe COVID-19 outcome | | | | | | | | | | | | | | | | |
| --- | --- | --- | --- | --- | --- | --- | --- | --- | --- | --- | --- | --- | --- | --- | --- | --- | --- | --- | --- | --- | --- | --- | --- |
|  |  |  |  |  |  |  | Hospitalization (non-intensive care unit) | | | | |  | Intensive care unit admission | | | | |  | Death | | | | |
|  | # of events | # of events | # of incident events | Cumulative  1-year incidence | Incidence rate |  | # of events | # of events | # of incident events | Cumulative  1-year incidence | Incidence rate |  | # of events | # of events | # of incident events | Cumulative  1-year incidence | Incidence rate |  | # of events | # of events | # of incident events | Cumulative  1-year incidence | Incidence rate |
|  | (total) | (in 1 year) | (in 1 year) | (per 10,000 persons) | (per 1,000 person-years) |  | (total) | (in 1 year) | (in 1 year) | (per 10,000 persons) | (per 1,000 person-years) |  | (total) | (in 1 year) | (in 1 year) | (per 10,000 persons) | (per 1,000 person-years) |  | (total) | (in 1 year) | (in 1 year) | (per 10,000 persons) | (per 1,000 person-years) |
| Overall | 6,191 | 8,476 | 7,532 | 28.5 | 2.2 |  | 4,789 | 6,530 | 6,442 | 24.4 | 1.9 |  | 432 | 550 | 550 | 2.1 | 0.2 |  | 970 | 1,396 | 1,396 | 5.3 | 0.4 |
| **Demographic characteristics** | | | |  |  |  |  |  |  |  |  |  |  |  |  |  |  |  |  |  |  |  |  |
| Age category (years) |  |  |  |  |  |  |  |  |  |  |  |  |  |  |  |  |  |  |  |  |  |  |  |
| 18-39 | 933 | 943 | 928 | 10.1 | 0.9 |  | 862 | 877 | 870 | 9.4 | 0.8 |  | 48 | 46 | 46 | 0.5 | 0.0 |  | 23 | 20 | 20 | 0.2 | 0.0 |
| 40-49 | 4492 | 517 | 485 | 10.1 | 0.9 |  | 366 | 414 | 405 | 8.4 | 0.7 |  | 58 | 71 | 71 | 1.5 | 0.1 |  | 25 | 32 | 32 | 0.7 | 0.1 |
| 50-59 | 733 | 898 | 825 | 18.1 | 1.6 |  | 578 | 714 | 698 | 15.3 | 1.4 |  | 81 | 98 | 98 | 2.2 | 0.2 |  | 74 | 86 | 86 | 1.9 | 0.2 |
| 60-64 | 534 | 671 | 604 | 26.5 | 2.8 |  | 400 | 503 | 493 | 21.6 | 2.3 |  | 68 | 82 | 82 | 3.6 | 0.4 |  | 66 | 86 | 86 | 3.8 | 0.4 |
| ≥65 | 3,542 | 5,447 | 4,690 | 84.7 | 8.1 |  | 2,583 | 4,022 | 3,976 | 71.8 | 6.8 |  | 177 | 253 | 253 | 4.6 | 0.5 |  | 782 | 1,172 | 1,172 | 21.2 | 2.1 |
| Sex |  |  |  |  |  |  |  |  |  |  |  |  |  |  |  |  |  |  |  |  |  |  |  |
| Female | 2,891 | 3,924 | 3,545 | 26.4 | 2.1 |  | 2,334 | 3,108 | 3,064 | 22.8 | 1.8 |  | 159 | 201 | 201 | 1.5 | 0.1 |  | 398 | 615 | 615 | 4.6 | 0.3 |
| Male | 3,300 | 4,552 | 3,987 | 30.8 | 2.3 |  | 2,455 | 3,422 | 3,378 | 26.1 | 1.9 |  | 273 | 349 | 349 | 2.7 | 0.2 |  | 572 | 781 | 781 | 6.0 | 0.5 |
| Other | 0 | 0 | 0 | 0.0 | 0.0 |  | 0 | 0 | 0 | 0.0 | 0.0 |  | 0 | 0 | 0 | 0.0 | 0.0 |  | 0 | 0 | 0 | 0.0 | 0.0 |
| Residence |  |  |  |  |  |  |  |  |  |  |  |  |  |  |  |  |  |  |  |  |  |  |  |
| Rural/urban |  |  |  |  |  |  |  |  |  |  |  |  |  |  |  |  |  |  |  |  |  |  |  |
| Rural | 1,132 | 1,431 | 1,265 | 40.7 | 3.2 |  | 841 | 1,057 | 1,049 | 33.8 | 2.6 |  | 107 | 125 | 125 | 4.0 | 0.3 |  | 184 | 249 | 249 | 8.0 | 0.6 |
| Urban | 5,059 | 7,045 | 6,267 | 26.9 | 2.1 |  | 3,948 | 5,473 | 5,393 | 23.2 | 1.8 |  | 325 | 425 | 425 | 1.8 | 0.1 |  | 786 | 1,147 | 1,147 | 4.9 | 0.4 |
| Long-term care |  |  |  |  |  |  |  |  |  |  |  |  |  |  |  |  |  |  |  |  |  |  |  |
| Yes | 143 | 271 | 241 | 175.8 | 16.2 |  | 49 | 94 | 92 | 67.1 | 6.1 |  | <10 | <10 | <10 | NA | NA |  | 92 | 172 | 172 | 125.4 | 11.7 |
| No | 6,048 | 8,205 | 7,291 | 27.8 | 2.1 |  | 4,740 | 6,436 | 6,350 | 24.2 | 1.9 |  | 423-431 | 541-549 | 541-549 | 2.1 | 0.2 |  | 878 | 1,224 | 1,224 | 4.7 | 0.4 |
| **Socioeconomic status** |  |  |  |  |  |  |  |  |  |  |  |  |  |  |  |  |  |  |  |  |  |  |  |
| Material deprivation index |  |  |  |  |  |  |  |  |  |  |  |  |  |  |  |  |  |  |  |  |  |  |  |
| 1 (most well off) | 799 | 1,209 | 1,077 | 21.3 | 1.6 |  | 633 | 930 | 915 | 18.1 | 1.4 |  | 44 | 64 | 64 | 1.3 | 0.1 |  | 122 | 215 | 215 | 4.2 | 0.3 |
| 2 | 802 | 1,200 | 1,079 | 21.6 | 1.6 |  | 605 | 926 | 919 | 18.4 | 1.3 |  | 69 | 82 | 82 | 1.6 | 0.2 |  | 128 | 192 | 192 | 3.8 | 0.3 |
| 3 | 1,051 | 1,449 | 1,287 | 25.9 | 2.0 |  | 806 | 1,114 | 1,105 | 22.3 | 1.7 |  | 72 | 97 | 97 | 2.0 | 0.2 |  | 173 | 238 | 238 | 4.8 | 0.4 |
| 4 | 1,254 | 1,713 | 1,519 | 29.5 | 2.2 |  | 961 | 1,332 | 1,308 | 25.4 | 1.9 |  | 81 | 103 | 103 | 2.0 | 0.2 |  | 212 | 278 | 278 | 5.4 | 0.4 |
| 5 (most deprived) | 1,776 | 2,098 | 1,860 | 37.5 | 3.1 |  | 1,374 | 1,605 | 1,584 | 32.0 | 2.6 |  | 143 | 165 | 165 | 3.3 | 0.3 |  | 259 | 328 | 328 | 6.6 | 0.5 |
| Missing | 509 | 807 | 710 | 55.8 | 3.9 |  | 410 | 623 | 611 | 48.1 | 3.5 |  | 23 | 39 | 39 | 3.1 | 0.2 |  | 76 | 145 | 145 | 11.4 | 0.7 |
| **Clinical characteristics** |  |  |  |  |  |  |  |  |  |  |  |  |  |  |  |  |  |  |  |  |  |  |  |
| Charlson Comorbidity Index |  |  |  |  |  |  |  |  |  |  |  |  |  |  |  |  |  |  |  |  |  |  |  |
| Category; burden |  |  |  |  |  |  |  |  |  |  |  |  |  |  |  |  |  |  |  |  |  |  |  |
| 0; none | 1,820 | 2,053 | 1,939 | 10.2 | 0.9 |  | 1,539 | 1,743 | 1,727 | 9.1 | 0.8 |  | 124 | 146 | 146 | 0.8 | 0.1 |  | 157 | 164 | 164 | 0.9 | 0.1 |
| 1-2; mild | 2,229 | 3,105 | 2,773 | 47.6 | 4.1 |  | 1,717 | 2,368 | 2,340 | 40.2 | 3.5 |  | 168 | 206 | 206 | 3.5 | 0.3 |  | 344 | 531 | 531 | 9.1 | 0.7 |
| 3-4; moderate | 1,169 | 1,805 | 1,555 | 151.5 | 13.8 |  | 841 | 1,326 | 1,304 | 127.1 | 11.5 |  | 84 | 113 | 113 | 11.0 | 1.2 |  | 244 | 366 | 366 | 35.7 | 3.4 |
| ≥5; severe | 973 | 1,513 | 1,265 | 267.2 | 24.9 |  | 692 | 1,093 | 1,071 | 226.2 | 21.0 |  | 56 | 85 | 85 | 18.0 | 1.7 |  | 225 | 335 | 335 | 70.8 | 7.0 |
| Health conditions of interest |  |  |  |  |  |  |  |  |  |  |  |  |  |  |  |  |  |  |  |  |  |  |  |
| Dialysis |  |  |  |  |  |  |  |  |  |  |  |  |  |  |  |  |  |  |  |  |  |  |  |
| Yes | 97 | 200 | 173 | 583.1 | 52.4 |  | 69 | 144 | 140 | 471.9 | 42.4 |  | 11 | 22 | 22 | 74.1 | 7.1 |  | 17 | 34 | 34 | 114.6 | 11.0 |
| No | 6,094 | 8,276 | 7,359 | 27.9 | 2.2 |  | 4,720 | 6,386 | 6,302 | 23.9 | 1.9 |  | 421 | 528 | 528 | 2.0 | 0.2 |  | 953 | 1,362 | 1,362 | 5.2 | 0.4 |
| Pulmonary hypertension |  |  |  |  |  |  |  |  |  |  |  |  |  |  |  |  |  |  |  |  |  |  |  |
| Yes | 61 | 97 | 83 | 360.2 | 33.5 |  | 46 | 71 | 69 | 299.5 | 28.3 |  | <10 | <10 | <10 | NA | NA |  | 10 | 17 | 17 | 73.8 | 6.4 |
| No | 6,130 | 8,379 | 7,449 | 28.2 | 2.2 |  | 4,743 | 6,459 | 6,373 | 24.2 | 1.9 |  | 423-431 | 423-431 | 423-431 | 2.1 | 0.2 |  | 960 | 1,379 | 1,379 | 5.2 | 0.4 |
| Dementia |  |  |  |  |  |  |  |  |  |  |  |  |  |  |  |  |  |  |  |  |  |  |  |
| Yes | 694 | 1,151 | 1,007 | 246.3 | 23.5 |  | 457 | 746 | 742 | 181.5 | 17.5 |  | 16 | 22 | 22 | 5.4 | 0.6 |  | 221 | 383 | 383 | 93.7 | 8.5 |
| No | 5,497 | 7,325 | 6,525 | 25.1 | 2.0 |  | 4,332 | 5,784 | 5,700 | 21.9 | 1.7 |  | 416 | 528 | 528 | 2.0 | 0.2 |  | 749 | 1,013 | 1,013 | 3.9 | 0.3 |
| History of a transplant |  |  |  |  |  |  |  |  |  |  |  |  |  |  |  |  |  |  |  |  |  |  |  |
| Yes | 163 | 392 | 323 | 294.0 | 18.7 |  | 111 | 274 | 261 | 237.6 | 15.2 |  | 26 | 57 | 57 | 51.9 | 3.7 |  | 26 | 61 | 61 | 55.5 | 3.7 |
| No | 6,028 | 8,084 | 7,209 | 27.4 | 2.1 |  | 4,678 | 6,256 | 6,181 | 23.5 | 1.8 |  | 406 | 493 | 493 | 1.9 | 0.2 |  | 944 | 1,335 | 1,335 | 5.1 | 0.4 |
| Parkinson's disease |  |  |  |  |  |  |  |  |  |  |  |  |  |  |  |  |  |  |  |  |  |  |  |
| Yes | 106 | 179 | 156 | 202.4 | 18.2 |  | 79 | 129 | 129 | 167.3 | 15.4 |  | <10 | <10 | <10 | NA | NA |  | 24 | 47 | 47 | 61.0 | 4.7 |
| No | 6,085 | 8,297 | 7,376 | 28.0 | 2.2 |  | 4,710 | 6,401 | 6,313 | 24.0 | 1.9 |  | 423-431 | 541-549 | 541-549 | 2.1 | 0.2 |  | 946 | 1,349 | 1,349 | 5.1 | 0.4 |
| Cancer |  |  |  |  |  |  |  |  |  |  |  |  |  |  |  |  |  |  |  |  |  |  |  |
| Yes | 482 | 840 | 694 | 166.6 | 14.1 |  | 332 | 610 | 589 | 141.4 | 11.7 |  | 38 | 59 | 59 | 14.2 | 1.4 |  | 112 | 171 | 171 | 41.0 | 4.1 |
| No | 5,785 | 7,636 | 6,838 | 26.3 | 2.1 |  | 4,457 | 5,920 | 5,853 | 22.5 | 1.8 |  | 394 | 491 | 491 | 1.9 | 0.2 |  | 858 | 1,225 | 1,225 | 4.7 | 0.3 |
| Chronic obstructive pulmonary disease | | | |  |  |  |  |  |  |  |  |  |  |  |  |  |  |  |  |  |  |  |  |
| Yes | 1,535 | 2,193 | 1,871 | 147.2 | 13.4 |  | 1,119 | 1,627 | 1,595 | 125.5 | 11.3 |  | 95 | 123 | 123 | 9.7 | 1.0 |  | 321 | 443 | 443 | 34.9 | 3.3 |
| No | 4,656 | 6,283 | 5,661 | 22.5 | 1.7 |  | 3,670 | 4,903 | 4,847 | 19.3 | 1.5 |  | 337 | 427 | 427 | 1.7 | 0.1 |  | 649 | 953 | 953 | 3.8 | 0.3 |
| Chronic kidney disease |  |  |  |  |  |  |  |  |  |  |  |  |  |  |  |  |  |  |  |  |  |  |  |
| Yes | 1,655 | 2,531 | 2,159 | 147.4 | 13.2 |  | 1,198 | 1,863 | 1,827 | 124.7 | 11.1 |  | 112 | 159 | 159 | 10.9 | 1.1 |  | 345 | 509 | 509 | 34.7 | 3.3 |
| No | 4,536 | 5,945 | 5,373 | 21.6 | 1.7 |  | 3,591 | 4,667 | 4,615 | 18.5 | 1.5 |  | 320 | 391 | 391 | 1.6 | 0.1 |  | 625 | 887 | 887 | 3.6 | 0.3 |
| Down's syndrome |  |  |  |  |  |  |  |  |  |  |  |  |  |  |  |  |  |  |  |  |  |  |  |
| Yes | 12 | 18 | 16 | 147.7 | 12.6 |  | <10 | 14 | 14 | 129.3 | 10.3 |  | <10 | <10 | <10 | NA | NA |  | <10 | <10 | <10 | NA | NA |
| No | 6,179 | 8,458 | 7,516 | 28.5 | 2.2 |  | 4,780-4,788 | 6,516 | 6,428 | 24.4 | 1.9 |  | 423-431 | 541-549 | 541-549 | 2.1 | 0.2 |  | 961-969 | 1,387-1,395 | 1,387-1,395 | 5.3 | 0.4 |
| Cardiovascular disease |  |  |  |  |  |  |  |  |  |  |  |  |  |  |  |  |  |  |  |  |  |  |  |
| Yes | 2,549 | 3,841 | 3,313 | 119.3 | 9.7 |  | 1,871 | 2,842 | 2,795 | 100.7 | 8.1 |  | 178 | 240 | 240 | 8.6 | 0.8 |  | 500 | 759 | 759 | 27.3 | 2.2 |
| No | 3,642 | 4,635 | 4,219 | 17.9 | 1.5 |  | 2,918 | 3,688 | 3,647 | 15.4 | 1.3 |  | 254 | 310 | 310 | 1.3 | 0.1 |  | 470 | 637 | 637 | 2.7 | 0.2 |
| Diabetes |  |  |  |  |  |  |  |  |  |  |  |  |  |  |  |  |  |  |  |  |  |  |  |
| Yes | 1,991 | 2,867 | 2,502 | 91.3 | 7.9 |  | 1,481 | 2,167 | 2,136 | 77.9 | 6.7 |  | 170 | 210 | 210 | 7.7 | 0.8 |  | 340 | 490 | 490 | 17.9 | 1.6 |
| No | 4,200 | 5,609 | 5,030 | 21.3 | 1.6 |  | 3,308 | 4,363 | 4,306 | 18.2 | 1.4 |  | 262 | 340 | 340 | 1.4 | 0.1 |  | 630 | 906 | 906 | 3.8 | 0.3 |
| Pregnancy |  |  |  |  |  |  |  |  |  |  |  |  |  |  |  |  |  |  |  |  |  |  |  |
| Yes | 50 | 55 | 55 | 88.2 | 7.6 |  | 50 | 55 | 55 | 88.2 | 7.6 |  | 0 | 0 | 0 | 0.0 | 0.0 |  | 0 | 0 | 0 | 0.0 | 0.0 |
| No | 6,141 | 8,421 | 7,477 | 28.4 | 2.2 |  | 4,739 | 6,475 | 6,387 | 24.3 | 1.9 |  | 432 | 550 | 550 | 2.1 | 0.2 |  | 970 | 1,396 | 1,396 | 5.3 | 0.4 |
| Other immunocompromising conditions | | | |  |  |  |  |  |  |  |  |  |  |  |  |  |  |  |  |  |  |  |  |
| Yes | 1,443 | 2,374 | 2,029 | 94.1 | 6.9 |  | 1,046 | 1,738 | 1,697 | 78.7 | 5.7 |  | 131 | 209 | 209 | 9.7 | 0.7 |  | 266 | 427 | 427 | 19.8 | 1.5 |
| No | 4,748 | 6,102 | 5,503 | 22.7 | 1.8 |  | 3,743 | 4,792 | 4,745 | 19.6 | 1.6 |  | 301 | 341 | 341 | 1.4 | 0.1 |  | 704 | 969 | 969 | 4.0 | 0.3 |
| Immunocompromised (overall) | | |  |  |  |  |  |  |  |  |  |  |  |  |  |  |  |  |  |  |  |  |  |
| Yes | 1,692 | 2,764 | 2,571 | 88.2 | 6.5 |  | 1,243 | 2,039 | 2,160 | 74.1 | 5.5 |  | 143 | 229 | 240 | 8.5 | 0.6 |  | 306 | 496 | 547 | 18.5 | 1.4 |
| No | 4,499 | 5,712 | 4,961 | 21.8 | 1.8 |  | 3,546 | 4,491 | 4,282 | 18.8 | 1.5 |  | 289 | 321 | 310 | 1.4 | 0.1 |  | 664 | 900 | 849 | 3.8 | 0.3 |
| Hypertension |  |  |  |  |  |  |  |  |  |  |  |  |  |  |  |  |  |  |  |  |  |  |  |
| Yes | 3,416 | 5,130 | 4,458 | 72.8 | 6.2 |  | 2,536 | 3,824 | 3,767 | 61.5 | 5.3 |  | 234 | 318 | 318 | 5.2 | 0.5 |  | 646 | 988 | 988 | 16.1 | 1.4 |
| No | 2,775 | 3,346 | 3,074 | 15.2 | 1.3 |  | 2,253 | 2,706 | 2,675 | 13.2 | 1.1 |  | 198 | 232 | 232 | 1.1 | 0.1 |  | 324 | 408 | 408 | 2.0 | 0.2 |
| Autoimmune disorders |  |  |  |  |  |  |  |  |  |  |  |  |  |  |  |  |  |  |  |  |  |  |  |
| Yes | 406 | 688 | 602 | 67.4 | 5.0 |  | 305 | 531 | 519 | 58.1 | 4.1 |  | 39 | 54 | 54 | 6.0 | 0.5 |  | 62 | 103 | 103 | 11.5 | 0.9 |
| No | 5,785 | 7,788 | 6,930 | 27.2 | 2.1 |  | 4,484 | 5,999 | 5,923 | 23.2 | 1.8 |  | 393 | 496 | 496 | 1.9 | 0.2 |  | 908 | 1,293 | 1,293 | 5.1 | 0.4 |
| Obesity |  |  |  |  |  |  |  |  |  |  |  |  |  |  |  |  |  |  |  |  |  |  |  |
| Yes | 1,054 | 1,398 | 1,247 | 54.6 | 4.6 |  | 800 | 1,063 | 1,051 | 46.0 | 3.8 |  | 105 | 130 | 130 | 5.7 | 0.5 |  | 149 | 205 | 205 | 9.0 | 0.7 |
| No | 5,137 | 7,078 | 6,285 | 26.1 | 2.0 |  | 3,989 | 5,467 | 5,391 | 22.4 | 1.7 |  | 327 | 420 | 420 | 1.7 | 0.1 |  | 821 | 1,191 | 1,191 | 4.9 | 0.4 |
| Mental health disorder | | |  |  |  |  |  |  |  |  |  |  |  |  |  |  |  |  |  |  |  |  |  |
| Yes | 1,705 | 2,261 | 2,042 | 50.0 | 4.0 |  | 1,357 | 1,754 | 1,731 | 42.3 | 3.4 |  | 123 | 155 | 155 | 3.8 | 0.3 |  | 225 | 352 | 352 | 8.6 | 0.6 |
| No | 4,486 | 6,215 | 5,490 | 24.6 | 1.9 |  | 3,432 | 4,776 | 4,711 | 21.1 | 1.6 |  | 309 | 395 | 395 | 1.8 | 0.1 |  | 745 | 1,044 | 1,044 | 4.7 | 0.4 |
| Asthma |  |  |  |  |  |  |  |  |  |  |  |  |  |  |  |  |  |  |  |  |  |  |  |
| Yes | 377 | 558 | 501 | 50.2 | 3.8 |  | 314 | 454 | 443 | 44.3 | 3.4 |  | 25 | 36 | 36 | 3.6 | 0.3 |  | 38 | 68 | 68 | 6.8 | 0.4 |
| No | 5,814 | 7,918 | 7,031 | 27.7 | 2.1 |  | 4,475 | 6,076 | 5,999 | 23.6 | 1.8 |  | 407 | 514 | 514 | 2.0 | 0.2 |  | 932 | 1,328 | 1,328 | 5.2 | 0.4 |
| Dyslipidemia |  |  |  |  |  |  |  |  |  |  |  |  |  |  |  |  |  |  |  |  |  |  |  |
| Yes | 1,596 | 2,268 | 1,998 | 30.9 | 2.6 |  | 1,219 | 1,751 | 1,718 | 26.6 | 2.2 |  | 136 | 166 | 166 | 2.6 | 0.3 |  | 241 | 351 | 351 | 5.4 | 0.4 |
| No | 4,595 | 6,208 | 5,534 | 27.8 | 2.1 |  | 3,570 | 4,779 | 4,724 | 23.7 | 1.8 |  | 296 | 384 | 384 | 1.9 | 0.2 |  | 729 | 1,045 | 1,045 | 5.2 | 0.4 |
| **Other factors** |  |  |  |  |  |  |  |  |  |  |  |  |  |  |  |  |  |  |  |  |  |  |  |
| Prior SARS-CoV-2 infection |  |  |  |  |  |  |  |  |  |  |  |  |  |  |  |  |  |  |  |  |  |  |  |
| Yes | 226 | 249 | 237 | 16.5 | 1.3 |  | 177 | 196 | 193 | 13.5 | 1.1 |  | 16 | 13 | 13 | 0.9 | 0.1 |  | 33 | 40 | 40 | 2.8 | 0.2 |
| No | 5,965 | 8,227 | 7,295 | 29.2 | 2.3 |  | 4,612 | 6,334 | 6,249 | 25.0 | 1.9 |  | 416 | 537 | 537 | 2.2 | 0.2 |  | 937 | 1,356 | 1,356 | 5.4 | 0.4 |
| Received nirmatrelvir/ritonavir | |  |  |  |  |  |  |  |  |  |  |  |  |  |  |  |  |  |  |  |  |  |  |
| Yes | 118 | 200 | 186 | 91.5 | 8.9 |  | 100 | 173 | 167 | 82.1 | 7.8 |  | <10 | 13 | 13 | 6.4 | 0.7 |  | <10 | 14 | 14 | 6.9 | 0.7 |
| No | 6,073 | 8,276 | 7,346 | 28.0 | 2.2 |  | 4,689 | 6,357 | 6,275 | 24.0 | 1.9 |  | 423-431 | 537 | 537 | 2.1 | 0.2 |  | 961-969 | 1,382 | 1,382 | 5.3 | 0.4 |
| Early vaccination |  |  |  |  |  |  |  |  |  |  |  |  |  |  |  |  |  |  |  |  |  |  |  |
| Yes | 5,629 | 7,952 | 7,054 | 28.7 | 2.2 |  | 4,351 | 6,119 | 6,037 | 24.6 | 1.9 |  | 385 | 504 | 504 | 2.1 | 0.2 |  | 893 | 1,329 | 1,329 | 5.4 | 0.4 |
| No | 562 | 524 | 478 | 25.7 | 2.2 |  | 438 | 411 | 405 | 21.8 | 1.9 |  | 47 | 46 | 46 | 2.5 | 0.2 |  | 77 | 67 | 67 | 3.6 | 0.3 |
| Pandemic wave vaccine dose received* | | | |  |  |  |  |  |  |  |  |  |  |  |  |  |  |  |  |  |  |  |  |
| Wave 2 | 55 | 91 | 80 | 51.8 | 3.6 |  | 20 | 37 | 37 | 24.0 | 1.5 |  | <10 | <10 | <10 | NA | NA |  | 34 | 52 | 52 | 33.7 | 2.6 |
| Wave 3 (Alpha) | 3,332 | 5,737 | 5,019 | 33.4 | 2.6 |  | 2,472 | 4,316 | 4,263 | 28.3 | 2.2 |  | 227-235 | 355-363 | 355-363 | 2.4 | 0.2 |  | 625 | 1,059 | 1,059 | 7.0 | 0.6 |
| Wave 4 (Delta) | 2,418 | 2,281 | 2,096 | 21.0 | 1.8 |  | 1,993 | 1,887 | 1,855 | 18.6 | 1.6 |  | 166 | 158 | 158 | 1.6 | 0.1 |  | 259 | 236 | 236 | 2.4 | 0.2 |
| Wave 5 (Omicron) | 386 | 367 | 337 | 27.8 | 2.5 |  | 304 | 290 | 287 | 23.7 | 2.2 |  | 30 | 28 | 28 | 2.3 | 0.2 |  | 52 | 49 | 49 | 4.0 | 0.4 |

*Pandemic wave was measured on the day follow-up commenced. Abbreviations: COVID-19 = coronavirus disease 2019; NA = not applicable; SARS-CoV-2 = severe acute respiratory syndrome coronavirus 2.

### Supplementary Table 5. Expected number of adults with a severe COVID-19 outcome (per 10,000 persons) after the index date among those who completed a primary vaccination series.

|  | Any severe COVID-19 outcome | | |  | Type of severe COVID-19 outcome | | | | | | | | | | | | | | |
| --- | --- | --- | --- | --- | --- | --- | --- | --- | --- | --- | --- | --- | --- | --- | --- | --- | --- | --- | --- |
|  |  |  |  |  | Hospitalization (non-intensive care unit) | | |  | | Intensive care unit admission | | | |  | | Death | | | |
|  | at 6-months | at 12-months | at 24-months |  | at 6-months | at 12-months | at 24-months | |  | | at 6-months | at 12-months | at 24-months | |  | | at 6-months | at 12-months | at 24-months |
|  | number  [95% CI] | number  [95% CI] | number  [95% CI] |  | number  [95% CI] | number  [95% CI] | number  [95% CI] | |  | | number  [95% CI] | number  [95% CI] | number  [95% CI] | |  | | number  [95% CI] | number  [95% CI] | number  [95% CI] |
| **Demographic characteristics** | |  |  |  |  |  |  | |  | |  |  |  | |  | |  |  |  |
| Age category (years) | |  |  |  |  |  |  | |  | |  |  |  | |  | |  |  |  |
| 18-39 | 3.6  [3.2-4.1] | 10.2  [9.4-11.0] | 14.7  [12.6-16.8] |  | 3.4  [3.0-3.8] | 9.6  [8.8-10.3] | 13.8  [11.7-15.9] | |  | | 0.2  [0.1-0.3] | 0.5  [0.3-0.7] | 0.8  [0.5-1.0] | |  | | 0.1  [0.0-0.1] | 0.2  [0.1-0.4] | 0.4  [0.2-0.5] |
| 40-49 | 3.8  [3.2-4.4] | 10.3  [9.1-11.4] | 13.8  [12.3-15.3] |  | 3.2  [2.7-3.8] | 8.6  [7.5-9.6] | 11.6  [10.3-13.0] | |  | | 0.5  [0.3-0.7] | 1.5  [1.1-2.0] | 2.1  [1.4-2.8] | |  | | 0.3  [0.1-0.4] | 0.6  [0.3-0.9] | 0.8  [0.5-1.2] |
| 50-59 | 5.4  [4.6-6.1] | 19.7  [17.9-21.6] | 32.3  [26.7-37.8] |  | 4.5  [3.8-5.2] | 16.9  [15.1-18.6] | 25.3  [22.6-28.1] | |  | | 0.6  [0.4-0.9] | 2.2  [1.6-2.8] | 5.9  [1.1-10.6] | |  | | 0.5  [0.3-0.7] | 1.8  [1.2-2.3] | 3.3  [2.5-4.2] |
| 60-64 | 7.4  [6.1-8.6] | 37.5  [33.2-41.9] | 64.2  [50.3-78.1] |  | 6.0  [4.8-7.1] | 31.1  [27.2-35.1] | 53.5  [39.8-67.1] | |  | | 1.0  [0.6-1.5] | 4.7  [3.2-6.2] | 8.3  [6.0-10.6] | |  | | 0.8  [0.4-1.2] | 5.3  [3.6-7.0] | 8.5  [6.1-10.8] |
| ≥65 | 20.6  [19.3-21.8] | 126.6  [120.0-133.2] | 243.6  [227.7-259.4] |  | 17.0  [15.8-18.2] | 109.9  [103.7-116.1] | 207.1  [192.4-221.7] | |  | | 1.6  [1.3-2.0] | 6.5  [5.1-7.8] | 11.6  [9.3-13.9] | |  | | 5.6  [4.9-6.3] | 32.8  [29.4-36.1] | 65.2  [57.9-72.6] |
| Sex |  |  |  |  |  |  |  | |  | |  |  |  | |  | |  |  |  |
| Female | 6.8  [6.3-7.3] | 24.5  [23.3-25.7] | 43.4  [39.6-47.2] |  | 5.9  [5.4-6.3] | 21.7  [20.6-22.8] | 37.3  [33.9-40.6] | |  | | 0.5  [0.3-0.6] | 1.3  [1.0-1.6] | 2.8  [1.8-3.9] | |  | | 1.0  [0.8-1.2] | 3.6  [3.1-4.0] | 7.1  [5.5-8.7] |
| Male | 8.6  [8.1-9.2] | 26.6  [25.4-27.8] | 46.8  [39.5-54.1] |  | 7.2  [6.7-7.7] | 22.6  [21.5-23.7] | 38.8  [32.4-45.3] | |  | | 0.9  [0.8-1.1] | 2.6  [2.2-3.0] | 3.6  [3.1-4.1] | |  | | 1.7  [1.5-1.9] | 5.1  [4.6-5.7] | 10.3  [6.9-13.6] |
| Residence |  |  |  |  |  |  |  | |  | |  |  |  | |  | |  |  |  |
| Rural | 12.3  [11.0-13.6] | 37.1  [34.3-39.9] | 58.6  [52.6-64.7] |  | 10.1  [8.9-11.3] | 30.7  [28.2-33.3] | 46.2  [42.6-49.9] | |  | | 1.5  [1.0-1.9] | 4.1  [3.1-5.0] | 7.0  [4.2-9.8] | |  | | 2.3  [1.7-2.9] | 7.4  [6.2-8.7] | 12.2  [8.0-16.3] |
| Urban | 7.1  [6.8-7.5] | 23.9  [23.0-24.7] | 42.7  [38.7-46.7] |  | 6.1  [5.7-6.4] | 20.9  [20.1-21.7] | 36.9  [33.1-40.6] | |  | | 0.6  [0.5-0.7] | 1.7  [1.4-1.9] | 2.6  [2.2-2.9] | |  | | 1.2  [1.1-1.4] | 3.9  [3.6-4.3] | 7.9  [6.3-9.4] |
| Long-term care/supportive living | | | |  |  |  |  | |  | |  |  |  | |  | |  |  |  |
| Yes | 43.7  [31.1-56.3] | 317.3  [196.4-436.7] | 1150.8  [788.8-1498.7] |  | 20.2  [11.5-28.8] | 92.3  [26.2-157.9] | 345.7  [153.1-534.4] | |  | | 0.0  [0.0-0.0] | 21.0  [0.0-62.1] | 21.0  [0.0-62.1] | |  | | 31.5  [20.8-42.3] | 239.6  [136.8-341.3] | 924.2  [589.4-1247.1] |
| No | 7.6  [7.2-7.9] | 25.1  [24.2-25.9] | 43.0  [39.8-46.2] |  | 6.5  [6.1-6.8] | 22.0  [21.2-22.8] | 37.2  [34.3-40.2] | |  | | 0.7  [0.6-0.8] | 1.9  [1.7-2.2] | 3.3  [2.6-4.0] | |  | | 1.2  [1.1-1.4] | 4.0  [3.6-4.3] | 7.2  [6.1-8.4] |
| **Socioeconomic status** | |  |  |  |  |  |  | |  | |  |  |  | |  | |  |  |  |
| Material Deprivation | |  |  |  |  |  |  | |  | |  |  |  | |  | |  |  |  |
| 1 | 5.7  [5.0-6.4] | 20.0  [18.3-21.8] | 39.0  [29.7-48.3] |  | 4.9  [4.3-5.6] | 17.5  [15.8-19.2] | 33.1  [24.2-41.9] | |  | | 0.4  [0.2-0.6] | 1.1  [0.7-1.5] | 1.7  [1.2-2.3] | |  | | 1.0  [0.7-1.3] | 3.2  [2.5-3.9] | 7.2  [4.2-10.3] |
| 2 | 5.9  [5.2-6.6] | 20.3  [18.6-22.1] | 31.6  [26.6-36.5] |  | 4.9  [4.3-5.5] | 17.0  [15.4-18.6] | 26.9  [22.0-31.8] | |  | | 0.6  [0.4-0.8] | 2.0  [1.5-2.5] | 2.5  [1.9-3.2] | |  | | 1.0  [0.7-1.3] | 3.5  [2.8-4.2] | 5.6  [4.1-7.1] |
| 3 | 7.1  [6.3-7.9] | 23.6  [21.8-25.4] | 46.2  [36.3-56.0] |  | 6.1  [5.4-6.8] | 20.6  [18.9-22.3] | 39.1  [30.8-47.5] | |  | | 0.7  [0.4-0.9] | 1.8  [1.3-2.3] | 2.5  [1.8-3.1] | |  | | 1.3  [1.0-1.7] | 4.3  [3.5-5.0] | 9.4  [4.1-14.6] |
| 4 | 7.7  [6.9-8.5] | 25.7  [23.9-27.6] | 44.2  [37.0-51.4] |  | 6.2  [5.5-6.9] | 22.6  [20.9-24.3] | 36.4  [30.5-42.3] | |  | | 0.8  [0.6-1.1] | 1.5  [1.1-1.9] | 2.8  [2.1-3.6] | |  | | 1.6  [1.2-1.9] | 4.6  [3.8-5.3] | 9.8  [5.7-13.9] |
| 5 | 12.1  [11.1-13.0] | 36.0  [34.0-38.1] | 58.2  [52.2-64.1] |  | 10.5  [9.6-11.4] | 31.2  [29.3-33.2] | 49.7  [44.3-55.1] | |  | | 1.0  [0.7-1.3] | 3.2  [2.5-3.8] | 6.0  [3.6-8.4] | |  | | 1.9  [1.5-2.3] | 5.8  [5.0-6.7] | 10.0  [8.4-11.6] |
| **Clinical characteristics** | | |  |  |  |  |  | |  | |  |  |  | |  | |  |  |  |
| Charlson Comorbidity Index Category; burden | | | | | |  |  | |  | |  |  |  | |  | |  |  |  |
| 0; none | 3.1  [2.8-3.4] | 10.2  [9.6-10.8] | 18.9  [15.8-22.0] |  | 2.7  [2.5-3.0] | 9.1  [8.6-9.7] | 17.2  [14.0-20.3] | |  | | 0.2  [0.2-0.3] | 0.7  [0.6-0.9] | 1.2  [0.9-1.4] | |  | | 0.3  [0.2-0.3] | 0.8  [0.7-1.0] | 1.7  [1.4-2.0] |
| 1-2; mild | 12.4  [11.5-13.4] | 51.5  [48.6-54.5] | 85.8  [78.6-93.0] |  | 10.3  [9.4-11.2] | 44.6  [41.9-47.4] | 71.5  [65.8-77.3] | |  | | 1.2  [0.9-1.5] | 4.0  [3.3-4.8] | 6.3  [5.2-7.4] | |  | | 2.1  [1.7-2.5] | 8.8  [7.6-10.0] | 17.3  [12.8-21.7] |
| 3-4; moderate | 42.1  [37.7-46.4] | 192.4  [176.1-208.7] | 332.4  [298.8-365.8] |  | 35.0  [31.0-38.9] | 161.4  [146.3-176.4] | 281.8  [249.6-313.9] | |  | | 4.4  [3.0-5.8] | 15.0  [10.6-19.3] | 22.4  [16.2-28.5] | |  | | 10.4  [8.2-12.6] | 46.6  [38.6-54.6] | 80.7  [66.9-94.5] |
| ≥5; severe | 76.3  [67.5-85.2] | 331.2  [299.7-362.7] | 605.5  [519.6-690.7] |  | 64.5  [56.3-72.7] | 284.7  [255.2-314.2] | 475.7  [416.9-534.2] | |  | | 5.9  [3.4-8.3] | 23.4  [15.1-31.7] | 58.9  [24.8-92.8] | |  | | 21.0  [16.3-25.7] | 90.9  [74.3-107.6] | 190.6  [132.8-248.0] |
| Health conditions of interest | |  |  |  |  |  |  | |  | |  |  |  | |  | |  |  |  |
| Dialysis |  |  |  |  |  |  |  | |  | |  |  |  | |  | |  |  |  |
| Yes | 202.9  [132.1-273.1] | 685.7  [473.1-893.6] | 1070.2  [771.0-1359.7] |  | 142.8  [81.8-203.5] | 576.7  [373.7-775.3] | 965.6  [670.8-1251.0] | |  | | 42.0  [10.7-73.2] | 69.3  [20.2-118.2] | 145.7  [0.0-301.3] | |  | | 37.1  [11.0-63.2] | 150.0  [54.5-244.5] | 207.3  [83.3-329.8] |
| No | 7.6  [7.2-7.9] | 25.2  [24.4-26.0] | 43.9  [40.5-47.2] |  | 6.4  [6.1-6.8] | 21.8  [21.1-22.6] | 37.0  [34.1-40.0] | |  | | 0.7  [0.6-0.8] | 1.9  [1.7-2.1] | 3.2  [2.5-3.9] | |  | | 1.3  [1.2-1.5] | 4.3  [3.9-4.6] | 8.4  [6.9-9.9] |
| Pulmonary hypertension | |  |  |  |  |  |  | |  | |  |  |  | |  | |  |  |  |
| Yes | 120.0  [68.0-171.7] | 522.1  [339.8-701.0] | 1087.7  [278.4-1829.7] |  | 90.1  [45.4-134.7] | 474.3  [297.0-648.4] | 988.6  [175.6-1734.4] | |  | | 19.0  [0.0-40.9] | 37.4  [0.0-79.5] | 65.5  [0.0-134.4] | |  | | 31.8  [6.3-57.1] | 71.4  [10.8-131.5] | 126.5  [29.0-223.1] |
| No | 7.6  [7.3-8.0] | 25.3  [24.4-26.1] | 43.6  [40.3-46.8] |  | 6.5  [6.1-6.8] | 21.9  [21.1-22.7] | 36.7  [33.9-39.6] | |  | | 0.7  [0.6-0.8] | 1.9  [1.7-2.2] | 3.2  [2.5-4.0] | |  | | 1.3  [1.2-1.5] | 4.3  [4.0-4.7] | 8.4  [6.9-9.9] |
| History of a transplant | |  |  |  |  |  |  | |  | |  |  |  | |  | |  |  |  |
| Yes | 85.9  [65.7-105.9] | 240.1  [187.6-292.4] | 403.1  [202.8-599.2] |  | 65.3  [47.6-82.8] | 197.0  [148.5-245.2] | 264.0  [201.1-326.5] | |  | | 20.7  [10.9-30.5] | 46.0  [24.6-67.4] | 140.8  [0.0-325.2] | |  | | 14.1  [6.4-21.8] | 37.4  [17.1-57.7] | 69.4  [33.4-105.2] |
| No | 7.4  [7.1-7.8] | 25.0  [24.1-25.8] | 43.3  [40.0-46.6] |  | 6.3  [6.0-6.6] | 21.6  [20.9-22.4] | 36.9  [33.9-39.8] | |  | | 0.6  [0.5-0.7] | 1.8  [1.6-2.0] | 2.8  [2.5-3.2] | |  | | 1.3  [1.2-1.5] | 4.3  [3.9-4.6] | 8.3  [6.8-9.8] |
| Dementia |  |  |  |  |  |  |  | |  | |  |  |  | |  | |  |  |  |
| Yes | 68.9  [59.8-78.0] | 356.7  [311.9-401.3] | 804.0  [693.7-912.9] |  | 53.7  [45.6-61.7] | 265.8  [226.6-304.8] | 560.2  [477.1-642.6] | |  | | 2.0  [0.4-3.6] | 15.2  [4.6-25.7] | 21.2  [7.8-34.6] | |  | | 23.3  [18.0-28.5] | 124.8  [98.8-150.7] | 340.5  [259.0-421.4] |
| No | 6.9  [6.5-7.2] | 23.1  [22.3-23.9] | 39.0  [36.0-42.0] |  | 5.9  [5.6-6.2] | 20.3  [19.6-21.1] | 34.2  [31.4-37.1] | |  | | 0.7  [0.6-0.8] | 1.9  [1.7-2.1] | 3.2  [2.5-3.9] | |  | | 1.1  [0.9-1.2] | 3.4  [3.1-3.7] | 5.8  [5.2-6.4] |
| Pregnancy |  |  |  |  |  |  |  | |  | |  |  |  | |  | |  |  |  |
| Yes | 62.5  [42.1-82.8] | 78.2  [54.2-102.1] | 85.1  [59.3-110.8] |  | 62.5  [42.1-82.8] | 78.2  [54.2-102.1] | 85.1  [59.3-110.8] | |  | | 0.0  [0.0-0.0] | 0.0  [0.0-0.0] | 0.0  [0.0-0.0] | |  | | 0.0  [0.0-0.0] | 0.0  [0.0-0.0] | 0.0  [0.0-0.0] |
| No | 7.6  [7.2-7.9] | 25.4  [24.6-26.2] | 44.2  [40.9-47.6] |  | 6.4  [6.1-6.7] | 22.0  [21.2-22.7] | 37.3  [34.3-40.3] | |  | | 0.7  [0.6-0.8] | 2.0  [1.7-2.2] | 3.3  [2.6-4.0] | |  | | 1.4  [1.2-1.5] | 4.4  [4.0-4.7] | 8.5  [7.0-10.0] |
| Cancer |  |  |  |  |  |  |  | |  | |  |  |  | |  | |  |  |  |
| Yes | 53.2  [45.3-61.0] | 181.9  [157.0-206.7] | 282.4  [246.5-318.3] |  | 43.3  [36.3-50.4] | 151.7  [128.9-174.5] | 233.7  [200.8-266.4] | |  | | 7.5  [4.5-10.4] | 14.5  [8.0-21.1] | 26.9  [15.5-38.2] | |  | | 15.4  [11.1-19.8] | 51.0  [37.7-64.3] | 83.3  [63.9-102.8] |
| No | 7.1  [6.7-7.4] | 24.0  [23.2-24.8] | 42.3  [38.9-45.7] |  | 6.0  [5.7-6.3] | 20.9  [20.1-21.6] | 35.8  [32.8-38.8] | |  | | 0.6  [0.5-0.7] | 1.8  [1.6-2.0] | 3.1  [2.4-3.8] | |  | | 1.2  [1.0-1.3] | 3.9  [3.6-4.2] | 7.9  [6.4-9.3] |
| Chronic obstructive pulmonary disease | | |  |  |  |  |  | |  | |  |  |  | |  | |  |  |  |
| Yes | 40.4  [36.6-44.2] | 168.6  [156.4-180.9] | 306.7  [273.7-339.6] |  | 34.4  [30.9-37.9] | 145.8  [134.3-157.3] | 255.5  [228.6-282.4] | |  | | 3.2  [2.1-4.2] | 12.0  [8.8-15.2] | 20.7  [15.7-25.6] | |  | | 10.3  [8.4-12.3] | 42.5  [36.3-48.7] | 81.1  [60.9-101.4] |
| No | 6.2  [5.8-6.5] | 20.4  [19.7-21.2] | 35.2  [32.1-38.2] |  | 5.2  [4.9-5.5] | 17.7  [17.0-18.4] | 29.9  [27.1-32.7] | |  | | 0.6  [0.5-0.7] | 1.6  [1.4-1.8] | 2.7  [2.0-3.5] | |  | | 0.9  [0.8-1.1] | 3.0  [2.7-3.3] | 5.9  [4.7-7.0] |
| Chronic kidney disease | |  |  |  |  |  |  | |  | |  |  |  | |  | |  |  |  |
| Yes | 39.5  [36.0-43.0] | 178.4  [165.6-191.1] | 319.7  [289.7-349.6] |  | 32.5  [29.3-35.7] | 151.1  [139.3-162.9] | 262.0  [238.7-285.2] | |  | | 3.8  [2.7-4.8] | 14.2  [10.7-17.7] | 28.1  [17.4-38.9] | |  | | 9.4  [7.7-11.2] | 43.9  [37.5-50.3] | 86.8  [68.3-105.1] |
| No | 6.0  [5.6-6.3] | 19.9  [19.2-20.7] | 34.4  [31.3-37.5] |  | 5.1  [4.8-5.4] | 17.4  [16.7-18.1] | 29.7  [26.8-32.6] | |  | | 0.5  [0.4-0.6] | 1.5  [1.3-1.7] | 2.3  [2.0-2.6] | |  | | 0.9  [0.8-1.0] | 2.9  [2.6-3.2] | 5.5  [4.4-6.6] |
| Parkinson's disease | |  |  |  |  |  |  | |  | |  |  |  | |  | |  |  |  |
| Yes | 38.6  [23.4-53.7] | 284.5  [195.3-372.9] | 918.5  [488.5-1329.1] |  | 27.9  [15.0-40.8] | 258.8  [172.2-344.7] | 785.3  [375.2-1177.9] | |  | | 1.7  [0.0-4.9] | 11.7  [0.0-31.6] | 27.2  [0.0-63.4] | |  | | 14.2  [4.9-23.6] | 49.8  [17.0-82.5] | 216.2  [53.7-376.1] |
| No | 7.6  [7.3-8.0] | 25.2  [24.3-26.0] | 42.5  [39.4-45.6] |  | 6.5  [6.1-6.8] | 21.8  [21.0-22.6] | 35.9  [33.2-38.5] | |  | | 0.7  [0.6-0.8] | 1.9  [1.7-2.2] | 3.3  [2.5-4.0] | |  | | 1.3  [1.2-1.5] | 4.3  [3.9-4.6] | 8.1  [6.7-9.5] |
| Cardiovascular disease | |  |  |  |  |  |  | |  | |  |  |  | |  | |  |  |  |
| Yes | 31.1  [28.9-33.4] | 119.5  [113.1-126.0] | 203.5  [185.4-221.5] |  | 25.8  [23.8-27.9] | 101.7  [95.7-107.7] | 165.4  [151.2-179.6] | |  | | 2.9  [2.3-3.6] | 9.5  [7.7-11.2] | 17.0  [11.6-22.3] | |  | | 7.6  [6.5-8.7] | 26.7  [23.7-29.7] | 49.9  [39.4-60.4] |
| No | 5.1  [4.8-5.4] | 16.9  [16.2-17.6] | 29.5  [26.6-32.4] |  | 4.4  [4.1-4.7] | 14.8  [14.2-15.5] | 25.9  [23.1-28.7] | |  | | 0.4  [0.4-0.5] | 1.2  [1.1-1.4] | 1.9  [1.7-2.2] | |  | | 0.7  [0.5-0.8] | 2.3  [2.0-2.5] | 4.4  [3.7-5.2] |
| Other immunocompromising conditions | | |  |  |  |  |  | |  | |  |  |  | |  | |  |  |  |
| Yes | 26.1  [23.8-28.5] | 85.6  [79.7-91.6] | 128.5  [113.9-143.0] |  | 20.8  [18.7-22.9] | 72.0  [66.5-77.5] | 105.7  [93.5-117.9] | |  | | 3.6  [2.8-4.5] | 8.5  [6.7-10.2] | 15.9  [8.3-23.5] | |  | | 6.4  [5.3-7.6] | 17.5  [14.8-20.1] | 24.8  [21.3-28.2] |
| No | 6.2  [5.8-6.5] | 21.1  [20.3-21.9] | 38.3  [34.9-41.7] |  | 5.3  [5.0-5.6] | 18.5  [17.7-19.2] | 32.6  [29.5-35.6] | |  | | 0.5  [0.4-0.5] | 1.5  [1.3-1.7] | 2.3  [2.0-2.6] | |  | | 0.9  [0.8-1.1] | 3.4  [3.1-3.7] | 7.3  [5.7-8.9] |
| Diabetes |  |  |  |  |  |  |  | |  | |  |  |  | |  | |  |  |  |
| Yes | 24.8  [22.8-26.8] | 105.1  [98.6-111.5] | 172.5  [156.1-188.9] |  | 20.6  [18.8-22.4] | 89.6  [83.6-95.6] | 140.5  [128.4-152.6] | |  | | 2.8  [2.1-3.4] | 9.3  [7.5-11.2] | 17.7  [11.6-23.8] | |  | | 4.6  [3.7-5.5] | 20.6  [17.7-23.4] | 38.4  [28.3-48.5] |
| No | 5.8  [5.4-6.1] | 18.8  [18.1-19.6] | 33.5  [30.2-36.7] |  | 4.9  [4.6-5.2] | 16.4  [15.7-17.1] | 28.9  [25.9-32.0] | |  | | 0.5  [0.4-0.6] | 1.3  [1.1-1.5] | 2.0  [1.7-2.3] | |  | | 1.0  [0.9-1.1] | 3.0  [2.7-3.3] | 5.8  [4.7-7.0] |
| Immunocompromised (overall) | | |  |  |  |  |  | |  | |  |  |  | |  | |  |  |  |
| Yes | 24.0  [22.0-25.9] | 79.9  [74.8-85.1] | 120.8  [108.6-133.1] |  | 19.2  [17.4-21.0] | 67.6  [62.8-72.4] | 100.5  [90.2-110.8] | |  | | 3.1  [2.4-3.8] | 7.4  [5.9-8.9] | 14.1  [7.7-20.5] | |  | | 5.7  [4.8-6.7] | 16.0  [13.7-18.2] | 23.7  [20.5-26.8] |
| No | 6.0  [5.6-6.3] | 20.5  [19.7-21.2] | 37.4  [33.9-40.9] |  | 5.2  [4.9-5.5] | 17.9  [17.1-18.6] | 31.8  [28.7-34.9] | |  | | 0.4  [0.4-0.5] | 1.4  [1.2-1.6] | 2.2  [1.9-2.5] | |  | | 0.9  [0.8-1.0] | 3.3  [2.9-3.6] | 7.1  [5.5-8.7] |
| Hypertension | |  |  |  |  |  |  | |  | |  |  |  | |  | |  |  |  |
| Yes | 18.4  [17.2-19.5] | 82.9  [78.9-86.9] | 142.5  [132.2-152.9] |  | 15.2  [14.2-16.3] | 71.3  [67.6-75.0] | 120.3  [111.3-129.3] | |  | | 1.8  [1.5-2.2] | 6.2  [5.2-7.2] | 10.9  [8.0-13.8] | |  | | 4.1  [3.6-4.7] | 18.2  [16.4-20.1] | 31.5  [26.7-36.3] |
| No | 4.6  [4.3-4.9] | 14.4  [13.7-15.0] | 25.2  [22.1-28.2] |  | 4.0  [3.7-4.3] | 12.6  [12.0-13.3] | 21.5  [18.8-24.2] | |  | | 0.4  [0.3-0.5] | 1.1  [0.9-1.3] | 1.7  [1.4-1.9] | |  | | 0.6  [0.4-0.7] | 1.6  [1.4-1.9] | 4.1  [2.7-5.4] |
| Autoimmune disorders | |  |  |  |  |  |  | |  | |  |  |  | |  | |  |  |  |
| Yes | 17.2  [14.2-20.1] | 60.7  [52.8-68.7] | 105.3  [73.8-136.7] |  | 12.8  [10.3-15.3] | 51.0  [43.6-58.3] | 92.9  [61.6-124.0] | |  | | 2.8  [1.6-4.0] | 6.5  [4.0-8.9] | 8.4  [5.3-11.4] | |  | | 4.1  [2.6-5.5] | 9.6  [6.6-12.7] | 15.0  [10.5-19.5] |
| No | 7.4  [7.0-7.7] | 24.5  [23.7-25.4] | 42.6  [39.3-46.0] |  | 6.3  [6.0-6.6] | 21.3  [20.5-22.1] | 35.9  [33.0-38.8] | |  | | 0.6  [0.5-0.7] | 1.8  [1.6-2.0] | 3.1  [2.4-3.9] | |  | | 1.3  [1.1-1.4] | 4.2  [3.9-4.5] | 8.3  [6.8-9.8] |
| Obesity |  |  |  |  |  |  |  | |  | |  |  |  | |  | |  |  |  |
| Yes | 15.8  [14.0-17.5] | 53.9  [49.6-58.2] | 89.9  [74.9-104.8] |  | 12.8  [11.2-14.3] | 46.0  [42.0-50.1] | 74.9  [61.8-88.0] | |  | | 2.0  [1.4-2.7] | 5.6  [4.3-7.0] | 12.0  [4.9-19.1] | |  | | 2.9  [2.2-3.7] | 8.5  [6.8-10.2] | 12.3  [9.8-14.8] |
| No | 7.0  [6.6-7.3] | 23.1  [22.2-23.9] | 40.4  [37.0-43.7] |  | 5.9  [5.6-6.3] | 20.0  [19.3-20.8] | 34.2  [31.2-37.2] | |  | | 0.6  [0.5-0.7] | 1.6  [1.4-1.8] | 2.5  [2.2-2.8] | |  | | 1.2  [1.1-1.4] | 4.0  [3.6-4.3] | 8.1  [6.5-9.8] |
| Mental health disorder | | |  |  |  |  |  | |  | |  |  |  | |  | |  |  |  |
| Yes | 13.9  [12.7-15.1] | 46.5  [43.7-49.4] | 87.1  [71.0-103.1] |  | 11.9  [10.8-13.0] | 40.4  [37.7-43.1] | 76.6  [60.9-92.3] | |  | | 1.2  [0.9-1.6] | 3.7  [2.9-4.5] | 5.3  [4.3-6.4] | |  | | 2.2  [1.7-2.7] | 6.3  [5.3-7.4] | 11.1  [8.0-14.3] |
| No | 6.6  [6.2-6.9] | 21.7  [20.8-22.5] | 36.8  [33.9-39.6] |  | 5.6  [5.2-5.9] | 18.7  [17.9-19.5] | 30.5  [28.2-32.8] | |  | | 0.6  [0.5-0.7] | 1.6  [1.4-1.8] | 2.9  [2.1-3.7] | |  | | 1.2  [1.1-1.4] | 4.0  [3.6-4.3] | 8.0  [6.3-9.6] |
| Down's syndrome | |  |  |  |  |  |  | |  | |  |  |  | |  | |  |  |  |
| Yes | 12.4  [0.0-36.7] | 201.4  [54.5-346.0] | 282.7  [97.9-464.0] |  | 12.4  [0.0-36.7] | 147.6  [23.6-269.9] | 229.3  [61.5-394.3] | |  | | 0.0  [0.0-0.0] | 17.4  [0.0-51.3] | 17.4  [0.0-51.3] | |  | | 0.0  [0.0-0.0] | 68.9  [0.0-163.8] | 68.9  [0.0-163.8] |
| No | 7.7  [7.4-8.1] | 25.5  [24.6-26.3] | 44.2  [40.9-47.6] |  | 6.5  [6.2-6.9] | 22.1  [21.3-22.8] | 37.4  [34.4-40.3] | |  | | 0.7  [0.6-0.8] | 1.9  [1.7-2.2] | 3.3  [2.6-4.0] | |  | | 1.4  [1.2-1.5] | 4.3  [4.0-4.7] | 8.5  [7.0-9.9] |
| Asthma |  |  |  |  |  |  |  | |  | |  |  |  | |  | |  |  |  |
| Yes | 12.2  [9.9-14.5] | 48.3  [42.0-54.6] | 67.3  [59.0-75.5] |  | 10.7  [8.5-12.8] | 43.5  [37.5-49.5] | 61.2  [53.2-69.1] | |  | | 1.1  [0.4-1.8] | 3.7  [2.0-5.4] | 5.0  [2.8-7.1] | |  | | 1.3  [0.6-2.1] | 5.8  [3.6-8.1] | 7.1  [4.5-9.6] |
| No | 7.5  [7.2-7.9] | 24.7  [23.9-25.5] | 43.5  [40.0-46.9] |  | 6.4  [6.0-6.7] | 21.3  [20.5-22.1] | 36.6  [33.5-39.6] | |  | | 0.7  [0.6-0.8] | 1.9  [1.7-2.1] | 3.2  [2.5-3.9] | |  | | 1.4  [1.2-1.5] | 4.3  [4.0-4.6] | 8.5  [7.0-10.0] |
| Dyslipidemia | |  |  |  |  |  |  | |  | |  |  |  | |  | |  |  |  |
| Yes | 8.2  [7.5-9.0] | 33.0  [30.8-35.2] | 55.4  [48.7-62.1] |  | 6.8  [6.1-7.4] | 28.7  [26.6-30.7] | 46.5  [41.4-51.6] | |  | | 1.0  [0.8-1.3] | 3.0  [2.3-3.6] | 4.4  [3.6-5.3] | |  | | 1.6  [1.2-1.9] | 5.4  [4.5-6.3] | 11.1  [6.6-15.5] |
| No | 7.6  [7.2-8.0] | 23.8  [22.9-24.6] | 41.8  [37.9-45.6] |  | 6.5  [6.1-6.8] | 20.6  [19.7-21.4] | 35.4  [31.9-38.9] | |  | | 0.6  [0.5-0.7] | 1.7  [1.5-1.9] | 3.0  [2.1-3.9] | |  | | 1.3  [1.1-1.5] | 4.1  [3.7-4.5] | 7.8  [6.4-9.2] |
| **Other factors** |  |  |  |  |  |  |  | |  | |  |  |  | |  | |  |  |  |
| Prior SARS-CoV-2 infection | |  |  |  |  |  |  | |  | |  |  |  | |  | |  |  |  |
| Yes | 7.3  [5.8-8.7] | 13.6  [11.4-15.8] | 29.7  [10.8-48.6] |  | 5.8  [4.5-7.1] | 11.1  [9.1-13.0] | 16.4  [13.6-19.1] | |  | | 0.4  [0.0-0.7] | 1.2  [0.5-1.8] | 2.0  [0.9-3.1] | |  | | 1.3  [0.7-1.9] | 2.0  [1.1-2.8] | 12.4  [0.0-31.1] |
| No | 7.7  [7.4-8.1] | 26.5  [25.6-27.4] | 45.5  [42.2-48.9] |  | 6.6  [6.2-6.9] | 23.0  [22.2-23.8] | 39.0  [35.8-42.1] | |  | | 0.7  [0.6-0.8] | 2.0  [1.8-2.2] | 3.4  [2.6-4.1] | |  | | 1.4  [1.2-1.5] | 4.5  [4.2-4.9] | 8.4  [7.1-9.6] |
| Received nirmatrelvir/ritonavir | | |  |  |  |  |  | |  | |  |  |  | |  | |  |  |  |
| Yes | 18.1  [11.5-24.7] | 149.2  [108.9-189.4] | 305.1  [230.7-378.8] |  | 15.2  [9.2-21.3] | 138.7  [99.1-178.1] | 274.1  [203.0-344.7] | |  | | 2.9  [0.3-5.4] | 6.6  [0.9-12.3] | 19.5  [0.0-39.0] | |  | | 0.6  [0.0-1.8] | 8.4  [0.0-17.9] | 36.0  [9.4-62.5] |
| No | 7.6  [7.3-8.0] | 25.1  [24.3-25.9] | 43.6  [40.2-46.9] |  | 6.5  [6.1-6.8] | 21.7  [20.9-22.5] | 36.7  [33.8-39.7] | |  | | 0.7  [0.6-0.8] | 1.9  [1.7-2.1] | 3.2  [2.5-3.9] | |  | | 1.4  [1.2-1.5] | 4.3  [4.0-4.7] | 8.4  [6.9-9.9] |
| Early vaccination | |  |  |  |  |  |  | |  | |  |  |  | |  | |  |  |  |
| Yes | 7.1  [6.8-7.5] | 26.4  [25.5-27.3] | 45.7  [42.3-49.0] |  | 6.0  [5.7-6.4] | 22.9  [22.1-23.8] | 38.6  [35.6-41.6] | |  | | 0.6  [0.5-0.7] | 2.0  [1.7-2.2] | 3.3  [2.6-4.0] | |  | | 1.3  [1.1-1.4] | 4.6  [4.2-5.0] | 8.8  [7.3-10.3] |
| No | 15.5  [13.7-17.4] | 24.1  [21.7-26.4] | 45.7  [42.3-49.0] |  | 13.1  [11.4-14.8] | 20.4  [18.3-22.6] | 38.6  [35.6-41.6] | |  | | 1.7  [1.1-2.3] | 2.4  [1.7-3.1] | 3.3  [2.6-4.0] | |  | | 2.2  [1.5-2.9] | 3.3  [2.5-4.2] | 8.8  [7.3-10.3] |
| Pandemic wave vaccine dose received* | | |  |  |  |  |  | |  | |  |  |  | |  | |  |  |  |
| Wave 2 | 9.5  [4.4-14.7] | 45.1  [21.2-68.9] | 138.5  [80.3-196.3] |  | 3.7  [0.5-6.9] | 27.5  [6.3-48.6] | 66.1  [26.0-106.1] | |  | | 0.0  [0.0-0.0] | 4.6  [0.0-13.7] | 4.6  [0.0-13.7] | |  | | 7.3  [2.8-11.9] | 21.2  [8.1-34.3] | 98.5  [48.6-148.1] |
| Wave 3 (Alpha) | 6.2  [5.8-6.6] | 38.3  [36.4-40.2] | 64.8  [60.7-68.8] |  | 5.1  [4.7-5.5] | 32.7  [30.9-34.4] | 54.9  [51.2-58.7] | |  | | 0.6  [0.5-0.8] | 3.1  [2.6-3.6] | 4.8  [3.9-5.8] | |  | | 1.5  [1.3-1.7] | 8.3  [7.4-9.2] | 13.7  [12.1-15.3] |
| Wave 4 (Delta) | 8.9  [8.3-9.5] | 20.2  [19.2-21.2] | 138.5  [80.3-196.3] |  | 7.8  [7.3-8.4] | 18.2  [17.2-19.1] | 66.1  [26.0-106.1] | |  | | 0.7  [0.5-0.8] | 1.3  [1.1-1.6] | 4.6  [0.0-13.7] | |  | | 1.0  [0.8-1.2] | 2.1  [1.8-2.4] | 98.5  [48.6-148.1] |
| Wave 5 (Omicron) | 16.1  [13.7-18.4] | 26.2  [23.2-29.2] | 64.8  [60.7-68.8] |  | 13.1  [11.0-15.2] | 22.2  [19.4-25.0] | 54.9  [51.2-58.7] | |  | | 1.7  [0.9-2.4] | 2.4  [1.5-3.4] | 4.8  [3.9-5.8] | |  | | 2.8  [1.8-3.7] | 3.9  [2.7-5.1] | 13.7  [12.1-15.3] |

*Pandemic wave was measured on the day follow-up commenced. Abbreviations: CI = confidence interval; COVID-19 = coronavirus disease 2019; SARS-CoV-2 = severe acute respiratory syndrome coronavirus 2.

### Supplementary Table 6. Unadjusted likelihood of a severe COVID-19 outcome among adults who completed a primary vaccination series, presented according to the type of severe COVID-19 outcome.

|  | Type of severe COVID-19 outcome | | |
| --- | --- | --- | --- |
|  | Hospitalization  (non-intensive care unit) | Intensive care unit admission | Death |
|  | unadjusted  hazard ratio [95% CI] | unadjusted  hazard ratio [95% CI] | unadjusted  hazard ratio [95% CI] |
| **Demographic characteristics** |  |  |  |
| Age (years) |  |  |  |
| Continuous (reference: one year lower) | 1.07 [1.06-1.07] | 1.05 [1.05-1.06] | 1.12 [1.12-1.13] |
| Category |  |  |  |
| 18-39 (reference) | - | - | - |
| 40-49 | 0.94 [0.83-1.07] | 2.68 [1.81-3.96] | 2.55 [1.44-4.53] |
| 50-59 | 1.88 [1.68-2.10] | 4.55 [3.15-6.57] | 8.84 [5.45-14.32] |
| 60-64 | 3.33 [2.94-3.77] | 9.70 [6.63-14.18] | 21.35 [13.11-34.77] |
| ≥65 | 11.37 [10.47-12.35] | 12.78 [9.18-17.80] | 138.49 [90.48-211.99] |
| Older age |  |  |  |
| ≥65 (reference: <65) | 8.61 [8.09-9.16] | 4.65 [3.79-5.69] | 33.21 [28.05-39.31] |
| Sex |  |  |  |
| Male (reference: female) | 1.03 [0.97-1.09] | 1.69 [1.39-2.07] | 1.43 [1.25-1.63] |
| Residence |  |  |  |
| Rural (reference: urban) | 1.49 [1.38-1.62] | 2.33 [1.86-2.92] | 1.66 [1.40-1.96] |
| Long-term care (reference: no) | 3.62 [2.65-4.94] | 0.85 [0.12-6.03] | 44.72 [35.56-56.24] |
| **Socioeconomic status** |  |  |  |
| Material deprivation index |  |  |  |
| 1 (most well off; reference) | - | - | - |
| 2 | 0.97 [0.87-1.09] | 0.73 [0.50-1.06] | 1.13 [0.86-1.49] |
| 3 | 1.12 [1.00-1.24] | 1.10 [0.79-1.52] | 1.54 [1.20-1.97] |
| 4 | 1.37 [1.24-1.51] | 1.23 [0.90-1.68] | 2.02 [1.60-2.54] |
| 5 (most deprived) | 1.75 [1.59-1.92] | 1.39 [1.03-1.88] | 2.05 [1.63-2.58] |
| **Clinical characteristics** |  |  |  |
| Charlson Comorbidity Index |  |  |  |
| Continuous (reference: one score lower) | 1.51 [1.50-1.52] | 1.49 [1.45-1.54] | 1.60 [1.58-1.63] |
| Category |  |  |  |
| 0; no comorbid burden (reference) | - | - | - |
| 1-2; mild comorbid burden | 4.69 [4.36-5.04] | 5.47 [4.30-6.95] | 9.02 [7.43-10.95] |
| 3-4; moderate comorbid burden | 17.05 [15.59-18.63] | 19.84 [14.89-26.45] | 47.25 [38.38-58.18] |
| ≥5; severe comorbid burden | 30.82 [27.98-33.94] | 32.18 [23.36-44.33] | 98.42 [79.60-121.71] |
| Health conditions of interest |  |  |  |
| (reference: not living with the condition) |  |  |  |
| Dialysis | 27.09 [20.80-35.27] | 51.56 [28.28-94.00] | 36.94 [22.52-60.62] |
| Pulmonary hypertension | 17.70 [13.01-24.07] | 21.39 [8.85-51.70] | 20.53 [11.00-38.29] |
| Dementia | 11.92 [10.75-13.22] | 3.96 [2.36-6.64] | 35.35 [30.10-41.53] |
| History of a transplant | 9.51 [7.80-11.60] | 25.71 [17.26-38.28] | 10.98 [7.32-16.48] |
| Parkinson's disease | 9.24 [7.28-11.73] | 3.89 [1.25-12.11] | 14.29 [9.35-21.84] |
| Chronic kidney disease | 8.38 [7.82-8.99] | 9.04 [7.25-11.27] | 14.06 [12.23-16.15] |
| Chronic obstructive pulmonary disease | 8.22 [7.65-8.83] | 7.33 [5.79-9.28] | 13.31 [11.56-15.32] |
| Cancer | 7.56 [6.71-8.51] | 9.94 [7.10-13.90] | 13.40 [10.90-16.47] |
| Cardiovascular disease | 6.74 [6.34-7.17] | 7.65 [6.28-9.32] | 11.47 [10.06-13.08] |
| Down's syndrome | 5.54 [2.77-11.08] | 7.03 [0.99-50.01] | 6.65 [1.66-26.65] |
| Hypertension | 5.32 [5.00-5.65] | 5.47 [4.49-6.65] | 9.54 [8.30-10.97] |
| Diabetes | 5.00 [4.68-5.33] | 7.15 [5.86-8.73] | 6.00 [5.22-6.89] |
| Other immunocompromising conditions | 3.81 [3.55-4.10] | 6.13 [4.97-7.56] | 5.17 [4.47-6.00] |
| Immunocompromised (overall) | 3.73 [3.48-3.99] | 5.49 [4.48-6.74] | 4.98 [4.32-5.73] |
| Pregnancy | 3.80 [2.82-5.12] | NA* | NA* |
| Autoimmune disorders | 2.39 [2.12-2.71] | 3.51 [2.52-4.91] | 2.47 [1.90-3.22] |
| Obesity | 2.25 [2.07-2.44] | 3.55 [2.82-4.45] | 1.99 [1.66-2.40] |
| Mental health disorder | 2.15 [2.01-2.30] | 2.16 [1.75-2.68] | 1.60 [1.36-1.87] |
| Asthma | 1.91 [1.69-2.16] | 1.77 [1.18-2.65] | 1.08 [0.77-1.53] |
| Dyslipidemia | 1.32 [1.23-1.41] | 1.76 [1.43-2.16] | 1.29 [1.11-1.51] |
| **Other factors** |  |  |  |
| Prior SARS-CoV-2 infection  (reference: no) | 0.53 [0.45-0.62] | 0.58 [0.35-0.96] | 0.48 [0.34-0.70] |

*The number of events were not sufficient to calculate the hazard ratio. Abbreviations: CI = confidence interval; COVID-19 = coronavirus disease 2019; NA = not applicable; SARS-CoV-2 = severe acute respiratory syndrome coronavirus 2.

### Supplementary Table 7. Unadjusted risk profiles among adults who completed a primary vaccination series.

|  | People who had a severe  COVID-19 outcome | Risk profile  population size | Hazard Ratio  Estimate | Population attributable risk |
| --- | --- | --- | --- | --- |
|  | n (%) | n (% of cohort) | unadjusted  hazard ratio [95% CI] | % |
| **Hospitalization (non-intensive care unit admission)** | *n=4,703* |  |  |  |
| Age ≥75 | 1,619 (34.4%) | 219,855 (8.3%) | 11.34 [10.65-12.08] | 28.5% |
| Age ≥65 | 2,509 (54.0%) | 553,854 (21.0%) | 8.60 [8.10-9.12] | 41.8% |
| Age ≥65, or hypertension | 3,163 (67.3%) | 846,335 (32.1%) | 7.10 [6.67-7.55] | 51.8% |
| Age ≥65, or CVD | 3,098 (65.9%) | 695,928 (26.4%) | 8.88 [8.35-9.44] | 53.7% |
| Age ≥65, or CVD, or COPD | 3,030 (64.4%) | 648,368 (24.6%) | 9.93 [9.34-10.56] | 52.8% |
| Age ≥65, or CVD, or diabetes | 3,367 (71.6%) | 815,744 (30.9%) | 8.82 [8.27-9.40] | 58.9% |
| Age ≥65, or CVD, or obesity | 3,319 (70.6%) | 840,815 (31.9%) | 7.67 [7.20-8.17] | 56.8% |
| Age ≥65, or CVD, or diabetes, or obesity | 3,514 (74.7%) | 933,539 (35.4%) | 7.92 [7.41-8.46] | 60.9% |
| Age ≥65, or any health condition of interest* | 4,703 (89.1%) | 1,442,900 (54.7%) | 8.90 [8.12-9.76] | 75.9% |
| CVD, or COPD | 2,285 (48.6%) | 361,096 (13.7%) | 7.58 [7.15-8.03] | 40.4% |
| Any health condition of interest* | 4,703 (86.2%) | 1,330,809 (50.4%) | 7.63 [7.02-8.29] | 72.2% |
| Immunocompromised status, or CKD | 2,044 (43.5%) | 414,626 (15.7%) | 5.30 [5.00-5.62] | 32.9% |
| Immunocompromised status, or CKD, or LTC | 2,067 (44.0%) | 422,279 (16.0%) | 5.33 [5.03-5.65] | 33.3% |
| **Intensive care unit admission** | *n=432* |  |  |  |
| Age ≥75 | 61 (14.1%) | 219,855 (8.3%) | 3.00 [2.28-3.96] | 6.3% |
| Age ≥65 | 177 (41.0%) | 553,854 (21.0%) | 4.54 [3.72-5.54] | 25.3% |
| Age ≥65, or hypertension | 282 (65.3%) | 846,335 (32.1%) | 6.16 [5.04-7.53] | 48.9% |
| Age ≥65, or CVD | 267 (61.8%) | 695,928 (26.4%) | 7.02 [5.76-8.55] | 48.1% |
| Age ≥65, or CVD, or COPD | 250 (57.9%) | 648,368 (24.6%) | 6.99 [5.75-8.50] | 44.2% |
| Age ≥65, or CVD, or diabetes | 316 (73.1%) | 815,744 (30.9%) | 9.11 [7.35-11.30] | 61.1% |
| Age ≥65, or CVD, or obesity | 293 (67.8%) | 840,815 (31.9%) | 6.45 [5.26-7.91] | 52.8% |
| Age ≥65, or CVD, or diabetes, or obesity | 331 (76.6%) | 933,539 (35.4%) | 8.45 [6.75-10.58] | 63.8% |
| Age ≥65, or any health condition of interest* | 432 (90.0%) | 1,442,900 (54.7%) | 9.62 [7.02-13.19] | 78.0% |
| CVD, or COPD | 211 (48.8%) | 361,096 (13.7%) | 7.45 [6.16-9.00] | 40.7% |
| Any health condition of interest* | 432 (88.2%) | 1,330,809 (50.4%) | 8.92 [6.65-11.95] | 76.2% |
| Immunocompromised status, or CKD | 206 (47.7%) | 414,626 (15.7%) | 6.13 [5.07-7.41] | 37.9% |
| Immunocompromised status, or CKD, or LTC | 207 (47.9%) | 422,279 (16.0%) | 6.09 [5.04-7.36] | 38.0% |
| **Death** | *n=970* |  |  |  |
| Age ≥75 | 570 (58.8%) | 846,335 (32.1%) | 33.72 [29.49-38.56] | 55.0% |
| Age ≥65 | 782 (80.6%) | 553,854 (21.0%) | 32.49 [27.62-38.22] | 75.5% |
| Age ≥65, or hypertension | 859 (88.6%) | 422,279 (16.0%) | 27.32 [22.39-33.33] | 83.2% |
| Age ≥65, or CVD | 855 (88.1%) | 695,928 (26.4%) | 35.03 [28.80-42.61] | 83.9% |
| Age ≥65, or CVD, or COPD | 841 (86.7%) | 361,096 (13.7%) | 36.96 [30.65-44.56] | 82.4% |
| Age ≥65, or CVD, or diabetes | 877 (90.4%) | 695,928 (26.4%) | 33.49 [27.02-41.51] | 86.1% |
| Age ≥65, or CVD, or obesity | 877 (90.4%) | 815,744 (30.9%) | 30.55 [24.65-37.86] | 85.9% |
| Age ≥65, or CVD, or diabetes, or obesity | 889 (91.6%) | 840,815 (31.9%) | 29.66 [23.61-37.26] | 87.1% |
| Age ≥65, or any health condition of interest* | 970 (96.4%) | 1,442,900 (54.7%) | 29.18 [20.82-40.90] | 92.0% |
| CVD, or COPD | 613 (63.2%) | 1,417,763 (53.7%) | 13.77 [12.08-15.70] | 57.4% |
| Any health condition of interest* | 970 (92.8%) | 1,330,809 (50.4%) | 15.66 [12.28-19.98] | 85.4% |
| Immunocompromised status, or CKD | 542 (55.9%) | 648,368 (24.6%) | 8.74 [7.69-9.92] | 47.7% |
| Immunocompromised status, or CKD, or LTC | 585 (60.3%) | 414,626 (15.7%) | 10.35 [9.10-11.78] | 52.8% |

*Health conditions of interest included asthma, cardiovascular disease, chronic kidney disease, chronic obstructive pulmonary disease, dementia, diabetes, Down's syndrome, hypertension, an immunocompromised status, a mental health disorder, obesity, Parkinson's disease, pregnancy, and pulmonary hypertension; dyslipidemia was not included. Hazard ratio estimates are unadjusted, with the reference group comprising individuals without the specified condition(s). Population attributable risk is unadjusted and based on observed incidence and risk factor prevalence in the cohort, estimating the proportion of severe COVID-19 outcomes potentially preventable by targeting these factors. Abbreviations: CI = confidence interval; CKD = chronic kidney disease; COPD = chronic obstructive pulmonary disease; COVID-19 = coronavirus disease 2019; CVD = cardiovascular disease; LTC = long-term care; NA = not applicable.
